# Outdoor air pollution and the risk of asthma exacerbations in single lag0 and lag1 exposure patterns: A systematic review and meta-analysis

**DOI:** 10.1101/2021.02.04.21251113

**Authors:** Junjun Huang, Xiaoyu Yang, Fangfang Fan, Yan Hu, Xi Wang, Sainan Zhu, Guanhua Ren, Guangfa Wang

## Abstract

**Background:** Asthma exacerbations accelerate the disease progression, as well as increases the incidence of hospitalizations and deaths. There have been studies on the effects of outdoor air pollution and asthma exacerbations; however, evidence regarding single lag0 and lag1 exposure patterns is inconclusive.

**Objective:** To synthesize evidence regarding the relationship between outdoor air pollution and the asthma exacerbation risk in single lag0 and lag1 exposure patterns.

**Methods:** We performed a systematic literature search using PubMed, Embase, Cochrane Library, Web of Science, ClinicalTrials, China National Knowledge Internet, Chinese BioMedical, and Wanfang databases until August 1^st^, 2020. Additionally, we reviewed the reference lists of the relevant articles. Two authors independently evaluated the eligible articles and performed structured extraction of relevant information. Pooled relative risks (RRs) and 95% confidence intervals (CIs) of lag0 and lag1 exposure patterns were estimated using the random-effect models.

**Results:** Eighty-four studies met the eligibility criteria and provided sufficient information for meta-analysis. Outdoor air pollutants were associated with significantly increased risks of asthma exacerbations in both single lag0 and lag1 exposure patterns [lag0: RR (95%CI) (pollutants), 1.057(1.011, 1.103) (air quality index; AQI), 1.007(1.005, 1.010) (PM_2.5_), 1.009(1.005, 1.012) (PM_10_), 1.010(1.006, 1.014) (NO_2_), 1.030(1.011, 1.048) (CO), 1.005(1.002, 1.009) (O_3_); lag1: RR (95%CI) (pollutants), 1.064(1.022, 1.106) (AQI), 1.005(1.002, 1.008) (PM_2.5_), 1.007(1.004, 1.011) (PM_10_), 1.008(1.004, 1.012) (NO_2_), 1.025(1.007, 1.042) (CO), 1.010(1.006, 1.013) (O_3_)], except SO_2_ [lag0: RR (95%CI), 1.004(1.000, 1.007); lag1: RR (95%CI), 1.003(0.999, 1.006)]. Subgroup analyses revealed stronger effects in children and asthma exacerbations associated with other events (including symptoms, lung function changes, and medication use as required).

**Conclusion:** These findings demonstrate that outdoor air pollution significantly increases the asthma exacerbation risk in single lag0 and lag1 exposure patterns.

**PROSPERO registration number:** CRD42020204097 (https://www.crd.york.ac.uk/PROSPERO).

**Strengths and limitations of this study:**

- We performed a systematic literature search of six databases (with no specified start date or language limitation).
- Secondary references were included.
- Publication bias was assessed by applying Begg’s and Egger’s tests.
- This study focused on the association between outdoor air pollution and the asthma exacerbation risk in single lag0 and lag1 exposure patterns.
- There were few available studies regarding the AQI, other events, and death analyses.

## INTRODUCTION

Asthma is a common chronic airway inflammatory disease that affects > 300 million people worldwide.[1] The global prevalence of clinical/treated asthma in adults is 4.5%; moreover, it varies among 70 countries by as much as 21-fold.[2] Epidemiologic studies have revealed an increasing asthma prevalence; moreover, asthma contributes to functional loss, increased healthcare costs, and severe medical complications.[3] Asthma exacerbations accelerate disease progression, as well as increase the incidence of hospitalizations and deaths.[4]

Outdoor air pollution has been made more and more attention to the serious consequences. Evidence has demonstrated that air pollution could cause critical public health problems. A retrospective study of 80, 515 deaths in Beijing during 2004-2008 has found that a reduction in life expectancy was associated with increased air pollution.[5] Several studies have shown that air pollution exposure increases the risk of asthma exacerbations.[6, 7, 8] However, the air pollution components are complex, including particulate matter diameter ≤ 2.5 μm (PM_2.5_), particulate matter diameter ≤ 10 μm (PM_10_), sulfur dioxide (SO_2_), carbon monoxide (CO), nitrogen dioxide (NO_2_), ozone (O_3_), etc. Martinez-Rivera et al. [6] reported that a positive association of the levels of NO_2_, but not SO_2_ and CO, with the number of accident and emergency room (ER) visits and hospitalizations for asthma exacerbations. However, another study reported a positive association of SO_2_, but not NO_2_, levels with pediatric asthma exacerbations. [7] Additionally, Ostro et al.[8] reported that new coughing episodes were associated with exposure to PM_10_, PM_2.5_, and NO_2_, but not O_3_. There have been inconsistent findings by studies on different pollutants.

A meta-analysis confirmed an association of the aforementioned six air pollutants with significantly increased risks of asthma emergency room visits and hospitalizations.[9] However, there are numerous pollutant types to be monitored; moreover, each pollutant has different effects on asthma exacerbations. Therefore, there is a need for a comprehensive pollution index that can represent the various pollutant effect and facilitate estimation of the air pollution level by the general public. The air quality index (AQI) is a useful comprehensive index that was adopted by the US Environmental Pollution Administration for daily air quality reporting to the general public in 1999.[10] The AQI includes sub-indices for PM_2.5_, PM_10_, SO_2_, NO_2_, CO, and O_3_.[11] Pan et al.[12] reported an association of AQI with an increased risk of hospitalizations for childhood asthma. Contrastingly, Letz et al.[13] reported no significant correlation of the AQI with the occurrence of ER visits for asthma in the basic military trainee population. There have been inconsistent findings regarding AQI and the asthma exacerbation risk. Additionally, multiple lag estimates, including single and cumulative lags, were selected in the overall analyses of the aforementioned meta-analysis. Lag exposure sensitivity analyses have employed different single lag patterns for different pollutants.[9] However, there is a need to understand whether pollutants contribute to asthma exacerbations on the same day or have lag effects. Moreover, other asthma exacerbation outcomes (e. g., symptoms) should be considered.

This systematic review and meta-analysis of time-series and case-crossover studies aimed to summarize existing evidence regarding the relationships of various pollutants (AQI, PM_2.5_, PM_10_, SO_2_, NO_2_, CO, and O_3_) with the asthma exacerbation risk (exacerbation-associated outpatient visits; ER visits; hospitalizations; deaths, and other events, including symptoms, lung function changes, and medication use as needed) in single lag0 and lag1 exposure patterns.

## METHODS

We follow the standardized program: Preferred Reporting Items for Systematic Reviews and Meta-Analyses guidelines.[14] There is no review protocol.

### Eligibility criteria and search strategy

The inclusion criteria were as follows: (1) a time-series or case-crossover study with original data; (2) asthma exacerbations associated with outpatient visits, ER visits, hospitalizations, deaths, or other events (symptoms, lung function changes, and medication use as needed) as outcomes; (3) inclusion of a study population of children, adults, or both; (4) using AQI, PM_2.5_, PM_10_, SO_2_, NO_2_, CO, or O_3_ as measurements for outdoor air pollution; and (5) reporting on the relationship between outdoor air pollution and asthma exacerbations.

The exclusion criteria were: (1) no single-pollutant model; (2) no data regarding single lag0 or lag1 exposure patterns; (3) having data that could not be recalculated into study-specific relative risks (RRs) and 95% confidence intervals (CIs) to a unit increase in AQI, a 1 mg/m^3^ increase in CO, and a 10μg/m^3^ in the other pollutants (PM_2.5_, PM_10_, SO_2_, NO_2_, and O_3_) by assuming a linear relationship of all pollutants.

We conducted a systematic literature search of PubMed, Embase, Cochrane Library, Web of Science, ClinicalTrials, China National Knowledge Internet, Chinese BioMedica, and Wanfang databases until August 1^st^, 2020 (no start date specified and no language limitation). We used a combination of keywords associated with the exposure types (e.g., “AQI”) and asthma exacerbation outcomes (e.g., “symptom increase”) (see supplementary Table S1 Search strategies for more details).

We excluded irrelevant articles by applying title screening. Articles meeting the inclusion criteria were included in the abstract reading. Moreover, we included the references of the articles that met the inclusion criteria. Among them, articles that did not meet the exclusion criteria were included in this systematic review and meta-analysis through full-text reading.

### Quality assessment and data extraction

Quality assessment was conducted based on Mustafic’s study with a maximum score of 5.[15] One point was assigned if asthma exacerbations were coded by the International Classification of Diseases, American Thoracic Society, National Asthma Education or Prevention Program, or International Classification of Primary Care 2. One point was assigned if pollutant measurements were performed daily with < 25% missing data. One point was assigned if long-term trends, seasonality, and temperature were all adjusted. One point was assigned if the humidity or day-of-week was adjusted. One point was assigned if influenza epidemics or holidays were adjusted.

Data extraction was performed using a standardized form, including the main characteristics (author, publication year, location, subgroup, study population, sample size, period, lag pattern, and quality score), outcome measures (asthma exacerbations related to outpatient visits; ER visits; hospitalizations; deaths; and other events, including symptoms, lung function changes, and medication use as needed), exposure measures (AQI, PM_2.5_, PM_10_, SO_2_, NO_2_, CO, and O_3_), and the study-specific RRs (95%CIs) of the single lag0 or lag1 exposure pattern. Eligible studies were examined and their relevant characteristics were independently recorded by two authors (XW and JH) in this standardized form. Any disagreements were resolved by consensus after discussion with an additional author (YH). When the same population was used in several publications, we only included the largest and most complete study.

### Statistical analysis and data synthesis

Standardized effect estimates were expressed as RRs and 95% CIs. The study-specific RRs were derived from single-pollutant models reporting RRs or percentage changes. We recalculated the study-specific RRs to a 100-unit, 1 mg/m^3^, 10μg/m^3^ increase in AQI, CO, and other pollutants (PM_2.5_, PM_10_, SO_2_, NO_2_, and O_3_), respectively, by assuming a linear relationship of all pollutants. We calculated the summary RRs of the single lag0 and lag1 exposure patterns. Heterogeneity was evaluated using the Q and I^2^-statistics through the random-effect model. An I^2^ statistic > 50%, 25%-50%, and < 25% indicates high, moderate, low heterogeneity, respectively.[16] Publication bias was assessed using Begg’s and Egger’s tests.[17, 18] To explore the heterogeneity in our pooled analysis, we applied sensitivity analyses based on studies with a quality of 4–5 scores. Various outcome analyses were conducted to combine the effects for evaluating differences in outpatient visits, ER visits, hospitalizations, deaths, and other events. Moreover, age-based subgroup analyses were conducted to evaluate differences between children (0–14 years) and adults (> 14 years). Statistical analyses were conducted using Stata/MP 14.0 (Stata, College Station, TX, USA). All tests were two-sided; moreover, statistical significance was defined as *P* < 0.05.

### Patient and public involvement

Given the nature of systematic review and meta-analysis, this study lacked patient or public involvement.

## RESULTS

### Study selection and eligible study characteristics

We initially identified 1169 articles. After reading the title, abstract, and full text, we included 84 articles. Figure 1 presents the approach to study selection.

Supplementary Table S2 presents the quality scores and main characteristics of the eligible studies.[12, 19-101] The outcomes were asthma exacerbations related to outpatient visits (9 studies), ER visits (44 studies), hospitalizations (29 studies), deaths (2 studies), and other events (7 studies). Additionally, one study did not specify separate data regarding ER visits and hospitalizations[96]; consequently, it was classified as ER visits or hospitalization outcomes. Regarding the single lag exposure pattern, 68 and 63 studies reported on lag0 and lag1, respectively. Regarding the age subgroups, 34 and 21 studies investigated children and adults, respectively.

### Overall and quality sensitivity analyses

In overall analyses, air pollutants were associated with significantly increased risks of asthma exacerbations in both of the single lag0 and lag1 exposure patterns [lag0: RR (95% CI) (pollutants), 1.057(1.011, 1.103) (AQI), 1.007(1.005, 1.010) (PM_2.5_), 1.009(1.005, 1.012) (PM_10_), 1.010(1.006, 1.014) (NO_2_), 1.030(1.011, 1.048) (CO), 1.005(1.002, 1.009) (O_3_); lag1: RR (95% CI) (pollutants), 1.064(1.022, 1.106) (AQI), 1.005(1.002, 1.008) (PM_2.5_), 1.007(1.004, 1.011) (PM_10_), 1.008(1.004, 1.012) (NO_2_), 1.025(1.007, 1.042) (CO), 1.010(1.006, 1.013) (O_3_)], except for SO_2_ [lag0: RR (95%CI), 1.004(1.000, 1.007); lag1: RR (95%CI), 1.003(0.999, 1.006)] (Table 1). The study-specific RRs presented high heterogeneity for all pollutants except for CO in the single lag1 exposure pattern. Table 1 shows the p values of Begg’s and Egger’s tests. There was no publication bias for AQI, PM_2.5_, NO_2_, and CO in the lag0 pattern, as well as AQI, PM_2.5_, SO_2_, and NO_2_ in the lag1 pattern. Supplementary Figure S1 shows the forest plot for the association between air pollutants and asthma exacerbations while Supplementary Figure S2 shows Begg’s funnel plot.

**Table 1.**
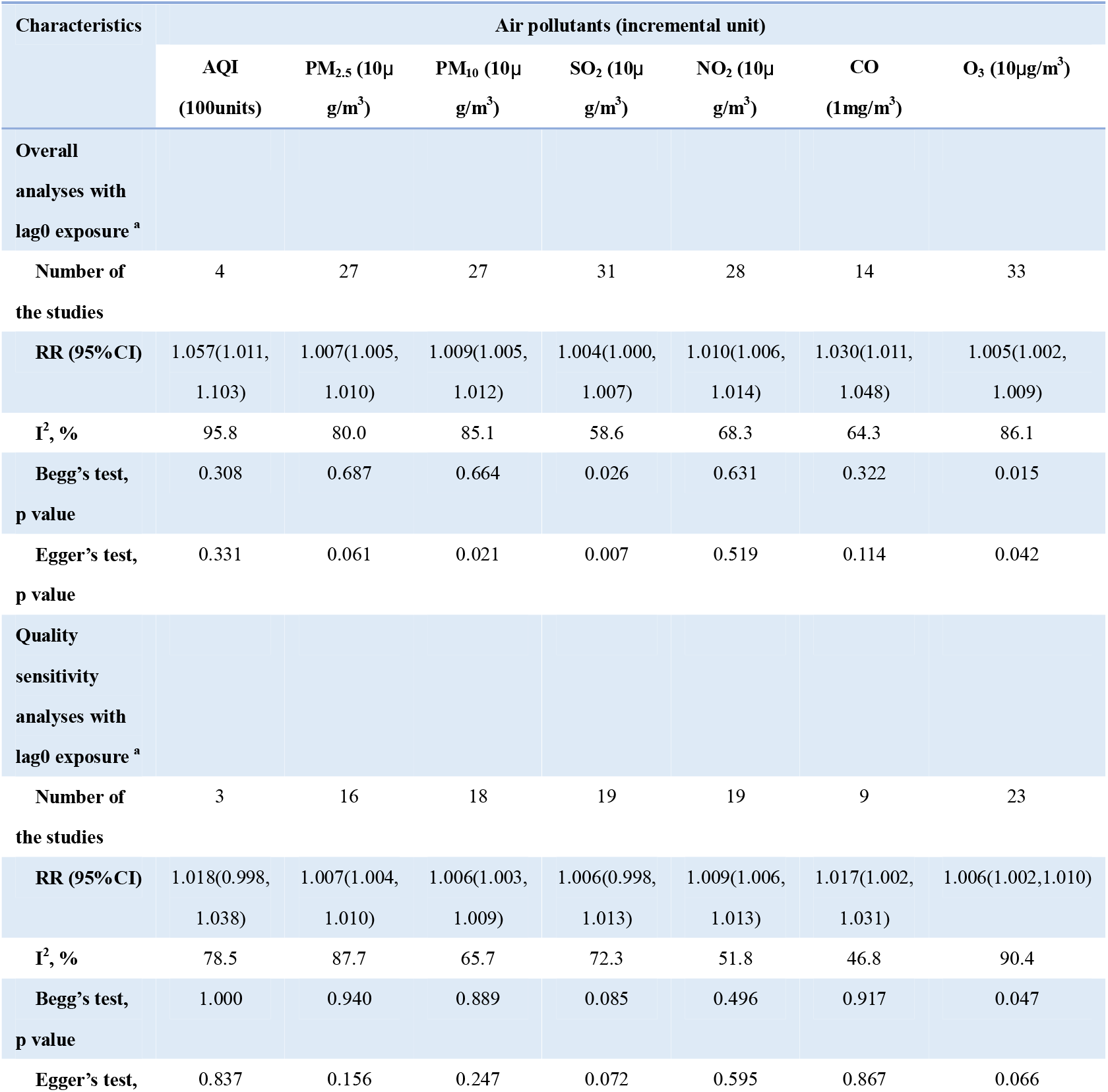
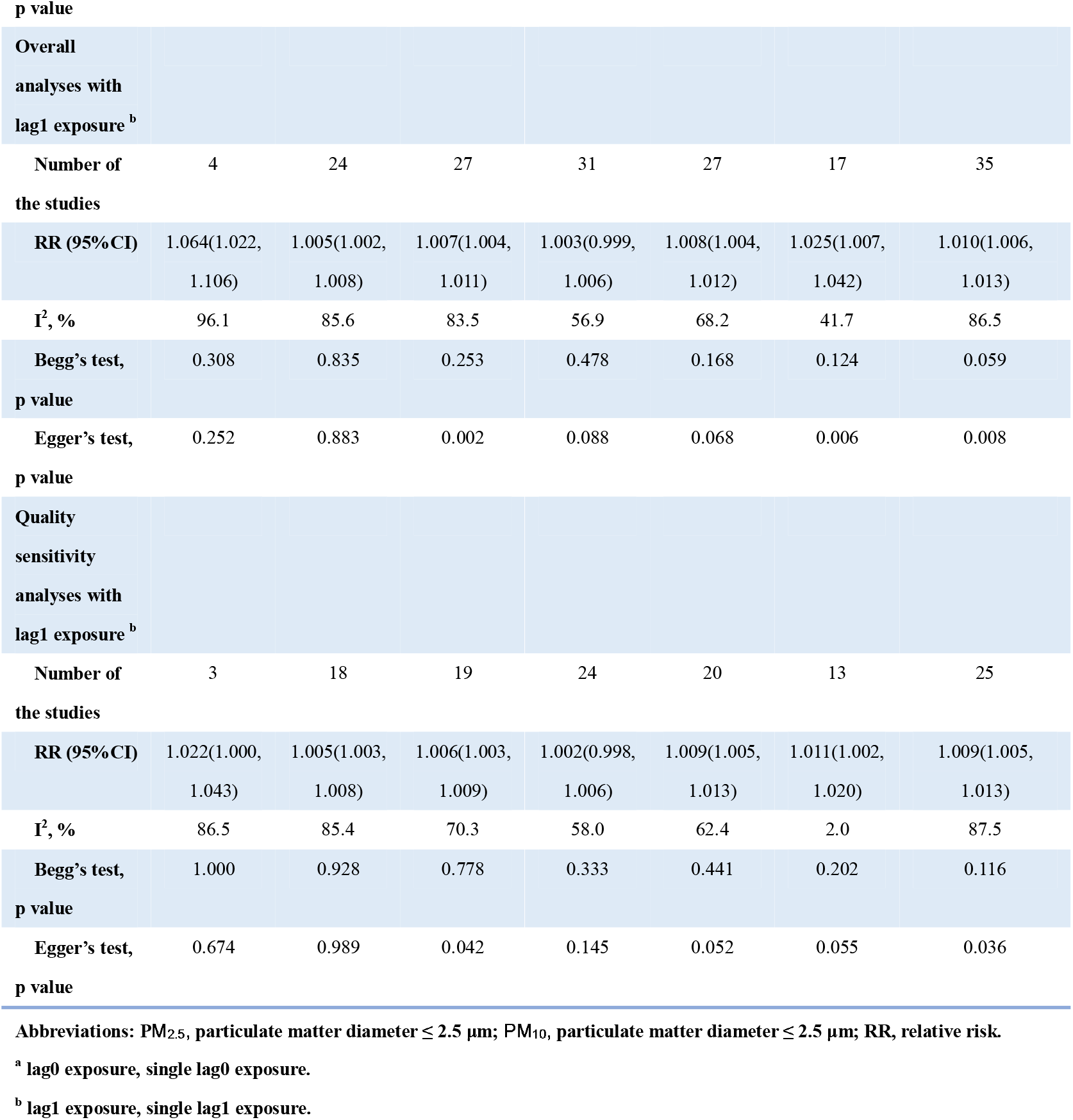
Relationships between air pollutants and asthma exacerbations in overall and quality sensitivity analyses.

Moreover, quality sensitivity analyses revealed a significant positive association of PM_2.5_, PM_10_, NO_2_, CO, and O_3_ with asthma exacerbations risks in both single lag0 and lag1 exposure patterns (Table 1). The study-specific RRs showed high heterogeneity for all pollutants except for CO. Publication bias was detected with Begg’s test for O_3_ in the lag0 pattern and Egger’s tests for PM_10_ and O_3_ in the lag1 pattern (Table 1). Supplementary Figure S3 shows the forest plot for relationships between air pollutants and asthma exacerbations while supplementary Figure S4 shows Begg’s funnel plot.

### Various outcome and age subgroup analyses

Various outcome analyses revealed more pronounced relationships for other event outcomes [lag0: RR (95% CI) (pollutants), 1.201(1.155, 1.247) (AQI), 1.047(1.024, 1.069) (PM_2.5_), 1.119(1.018, 1.220) (PM_10_), 1.033(1.000, 1.066) (NO_2_), 1.124(1.051, 1.197) (CO); lag1: RR (95% CI) (pollutants), 1.204(1.158, 1.251) (AQI), 1.080(1.010, 1.150) (PM_2.5_), 1.122(1.015, 1.230) (PM_10_), 1.046(1.013, 1.079) (NO_2_), 1.155(1.037, 1.274) (CO)], except for SO_2_ and O_3_. Supplementary Table S3 presents details regarding the relationship between air pollutants and asthma exacerbations in various outcomes analyses. Supplementary Figure S1 shows the forest plot.

Age-based subgroup analyses revealed stronger relations in children than in adults, except for NO_2_ in the lag1 pattern and O_3_ in both patterns. There was no tendency toward a stronger relation for AQI given the AQI adult subgroup lacked eligible studies. Supplementary Table S3 presents the details regarding the relationship between air pollutants and asthma exacerbations in various outcomes and age subgroup analyses. Figure 2 and 3 shows the forest plot.

## DISCUSSION

### Summary of evidence

Our study provides novel evidence that air pollution exposure significantly increases the asthma exacerbation risk; specifically, AQI, PM_2.5_, PM_10_, NO_2_, CO, and O_3_ contribute to asthma exacerbations with single lag0 and lag1 exposure patterns. This suggests that these pollutants cause asthma exacerbations on the day of air pollution onset. Moreover, the AQI may be a good index representing the asthma exacerbation risk during air pollution. Mechanisms underlying the association of air pollution with asthma exacerbations include stimulation of airway epithelium and inflammatory cells, oxidative stress responses, respiratory cells, respiratory reflex responses, and epigenetic modifications.[102] Exposure to PM modulates the airway epithelium and promotes the production of several cytokines, including IL-1, IL-6, IL-8, IL-25, IL-33, TNF-α, and GM-CSF.[103] In a mouse model, there was an ozone-induced increase in bronchoalveolar lavage (BAL) levels of IL-23, which is an important cytokine for IL-17A+ cell recruitment and activation, at 24 h after ozone exposure (2 ppm for 3 h).[104] Willart et al.[105] reported an increase in lung IL-1α levels in naïve mice at 2 h and 24 h after exposure. Another study reported an increase in IL-1 αlevels within the first 24 h after ozone exposure (1 ppm for 1 h).[106] A study assessed sensitized mice before (0 h) and 1, 6, 12, 24, and 72 h after exposure and reported that the 12- and 24-h group had the highest cytokine levels and cell counts in BAL fluid.[107] Diesel exhaust particles were found to induce an increase in IL-17A levels in primary bronchial epithelial cells of patients with asthma at 2 post-exposure hours.[108] Moreover, Zhang et al.[109] reported that diesel exhaust particles induced increased *TET1* expression in human bronchial epithelial cells at 1 post-exposure hour. Therefore, in case air pollution exposure cannot be avoided, patients with asthma should employ adequate strategies (including asthma medications) to reduce damage on the same day.

In lag0 and lag1 exposure patterns, we found that the RRs of air pollution and asthma exacerbations related to other events (including symptoms, lung function changes, and medication use as needed) were much higher than those of outpatient visits, ER visits, hospitalizations, and deaths. There has been no study on the relationship between outdoor air pollution and other events, as well as outpatient visits, ER visits, hospitalizations, or deaths within the same population. However, a prospective case-control study on children reported an association of AQI, PM_2.5_, NO_2_, and O_3_ with increased expression of multiple inflammatory airway epithelial responses, as well as a decline in FEV_1_%predicted in the lead up to clinical asthma exacerbations on the exacerbation-onset day without a viral trigger.[110] Pan et al.[12] reported a positive correlation between AQI and childhood asthma hospitalizations that appeared and peaked on lag3 and lag9 days, respectively. Additionally, there are positive correlations of asthma deaths with PM_2.5_[32] on lag3 day or SO_2_ and NO_2_^79^ on lag2 day. This could be attributed to changes in symptoms and lung function being the initial manifestations of airway inflammation caused by air pollution. Subsequently, patients use relief medications for these changes and if they do not improve, they seek medical help and are recorded as outpatient visits, ER visits, or hospitalizations. Therefore, when patients with asthma present with symptoms or lung function changes during air pollution, they should receive active treatment to block asthma exacerbation progression.

It has been confirmed that children with asthma were more susceptible to outdoor air pollution. This could be attributed to immature lung growth and host-defense capacity in children.[9] Since children spend more time outdoors than adults, they inhale more air pollutants per pound of body weight.[111] Additionally, children may have low symptom tolerance and parents may be more active in taking their children to hospitals.

### Validity of results

This study has several strengths, including identification of potential articles based on a systematic literature search of six databases (no start date specified and no language limitation) and inclusion of secondary references. Furthermore, two authors examined the eligible studies based on the inclusion and exclusion criteria followed by independent data extraction using a priori set method in the standardized form. Finally, any disagreements were discussed together with a third author.

### Limitations

This study has several limitations. First, there were few available studies for the AQI, other events, and death analyses. Therefore, findings regarding the AQI, deaths, and other events analyses should be cautiously interpreted. Second, there was a high heterogeneity degree in most analyses. This might be associated with varying study outcomes and design qualities. Nonetheless, we reduced the heterogeneity degrees in quality sensitivity and various outcome analyses.

## CONCLUSIONS

This systematic review and meta-analysis provides new evidence indicating that air pollution exposure significantly increases the asthma exacerbation risk. Specifically, AQI, PM_2.5_, PM_10_, NO_2_, CO, and O_3_ contribute to asthma exacerbations with single lag0 and lag1 exposure patterns. In case air pollution exposure cannot be avoided, patients with asthma should employ adequate strategies (including asthma medications) to reduce same-day damage.

## Supporting information

Figure S1 Forest plot for relationships between air pollutants and asthma exacerbations in overall and various outcomes analyses

Figure S2. Begg's funnel plot for relationships between air pollutants and asthma exacerbations in overall analyses

Figure S3 Forest plot for relationships between air pollutants and asthma exacerbations in quality sensitivity analyses

Figure S4. Begg's funnel plot for relationships between air pollutants and asthma exacerbations in quality sensitivity analyses

PRISMA 2009 checklist(for asthma meta)

Table S1. Search strategies

Table S2. The quality scores and the main characteristics of the eligible studies

Table S3. Relationships between air pollutants and asthma exacerbations in various outcomes and age subgroup analyses

## Data Availability

Data are available on reasonable request.

## ACKNOWLEDGEMENTS

The authors thank Dr. Pengkang He from Peking University First Hospital for his help.

## CONTRIBUTORS

JH and XY are joint first authors. JH obtained funding. YH and GW are joint corresponding authors. JH, YH, and GW conceived and designed the study. JH and XY performed a systematic literature search under the supervision of GR and YH. JH, YH, and XW reviewed the articles and performed data extraction. JH and FF analyzed the data under the supervision of SZ, YH, and GW. JH, YH, XY, and FF wrote the manuscript. All authors read and approved the final manuscript and were responsible for their contributions.

## DATA AVAILABILITY STATEMENT

Data are available on reasonable request.

---

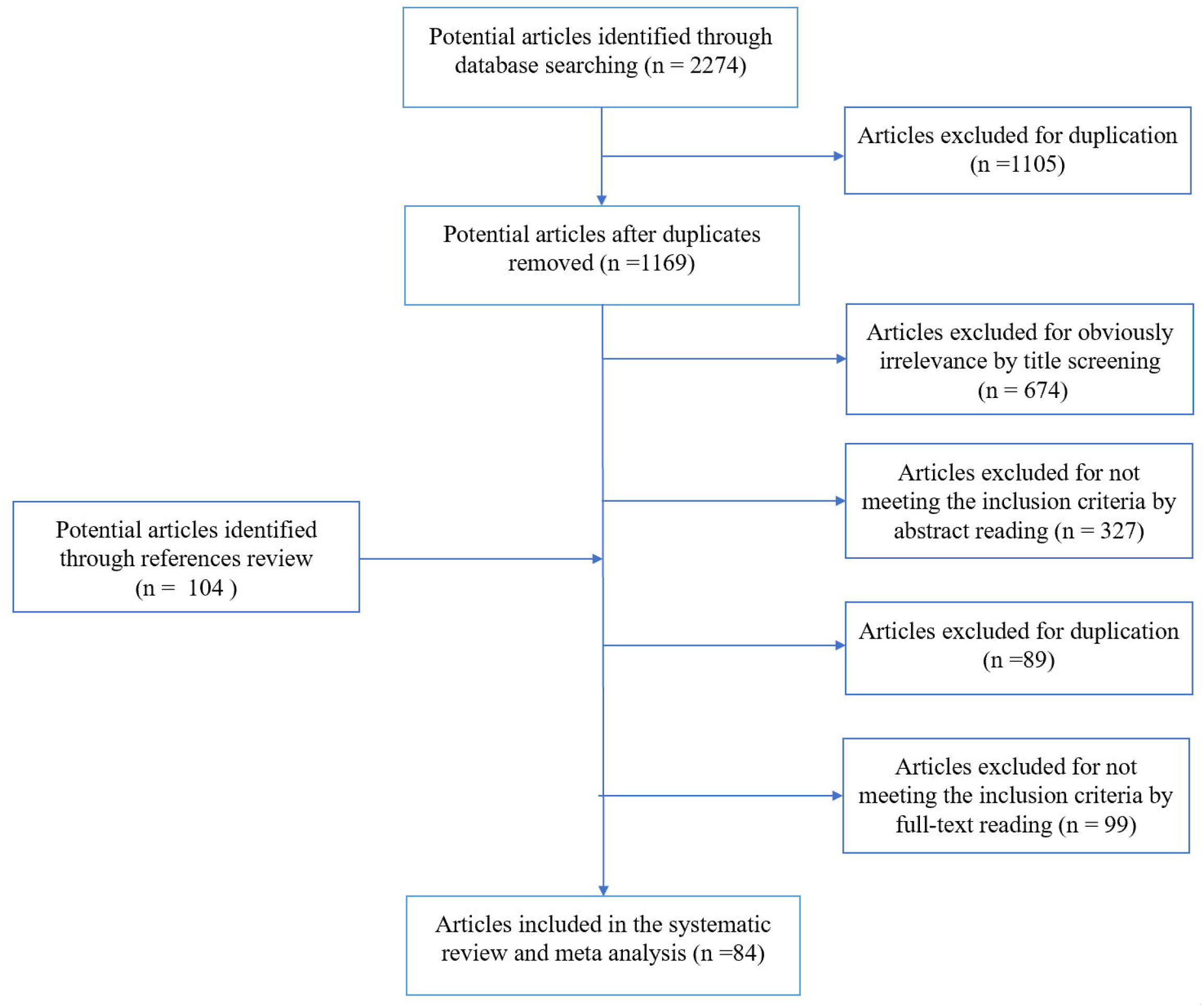

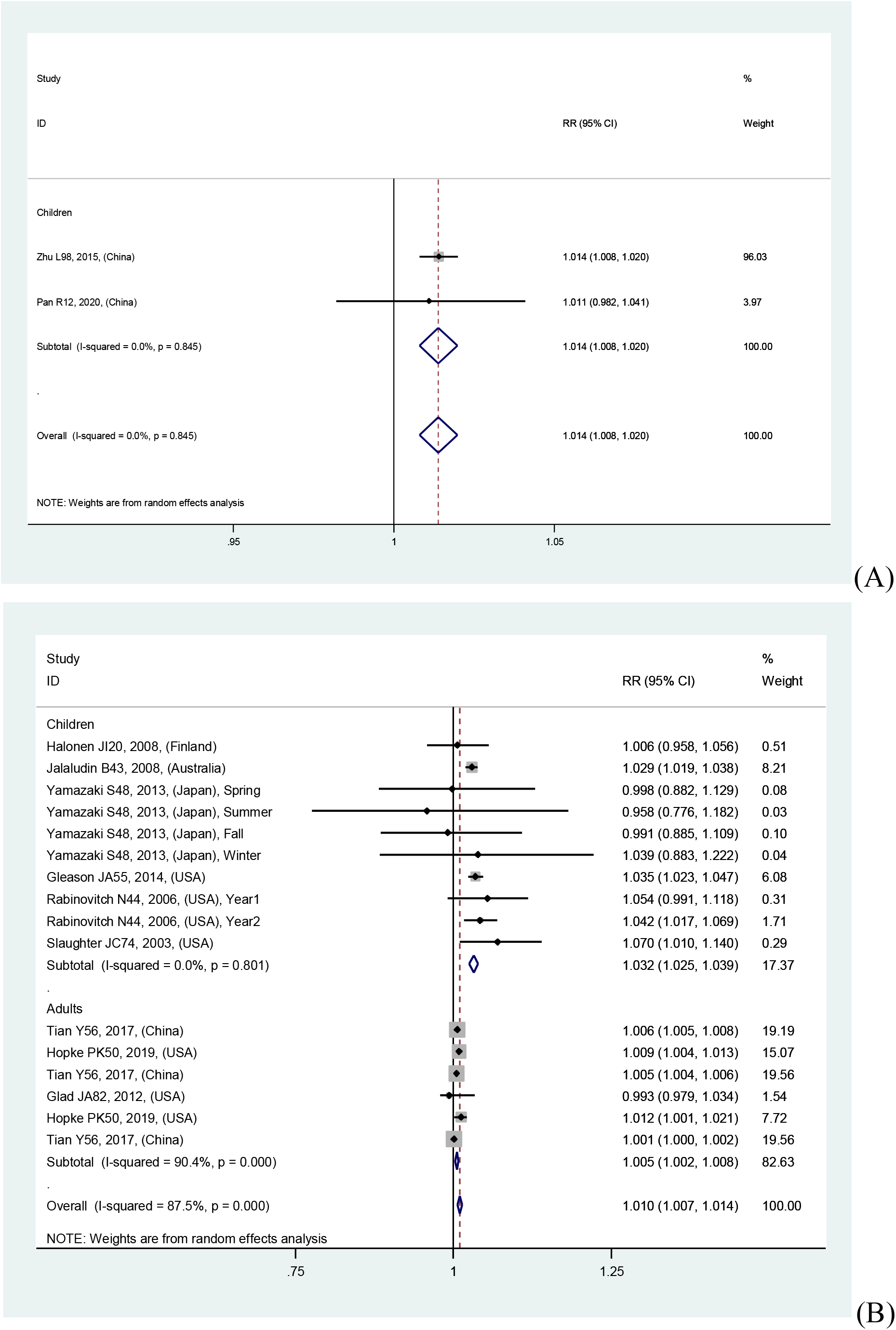
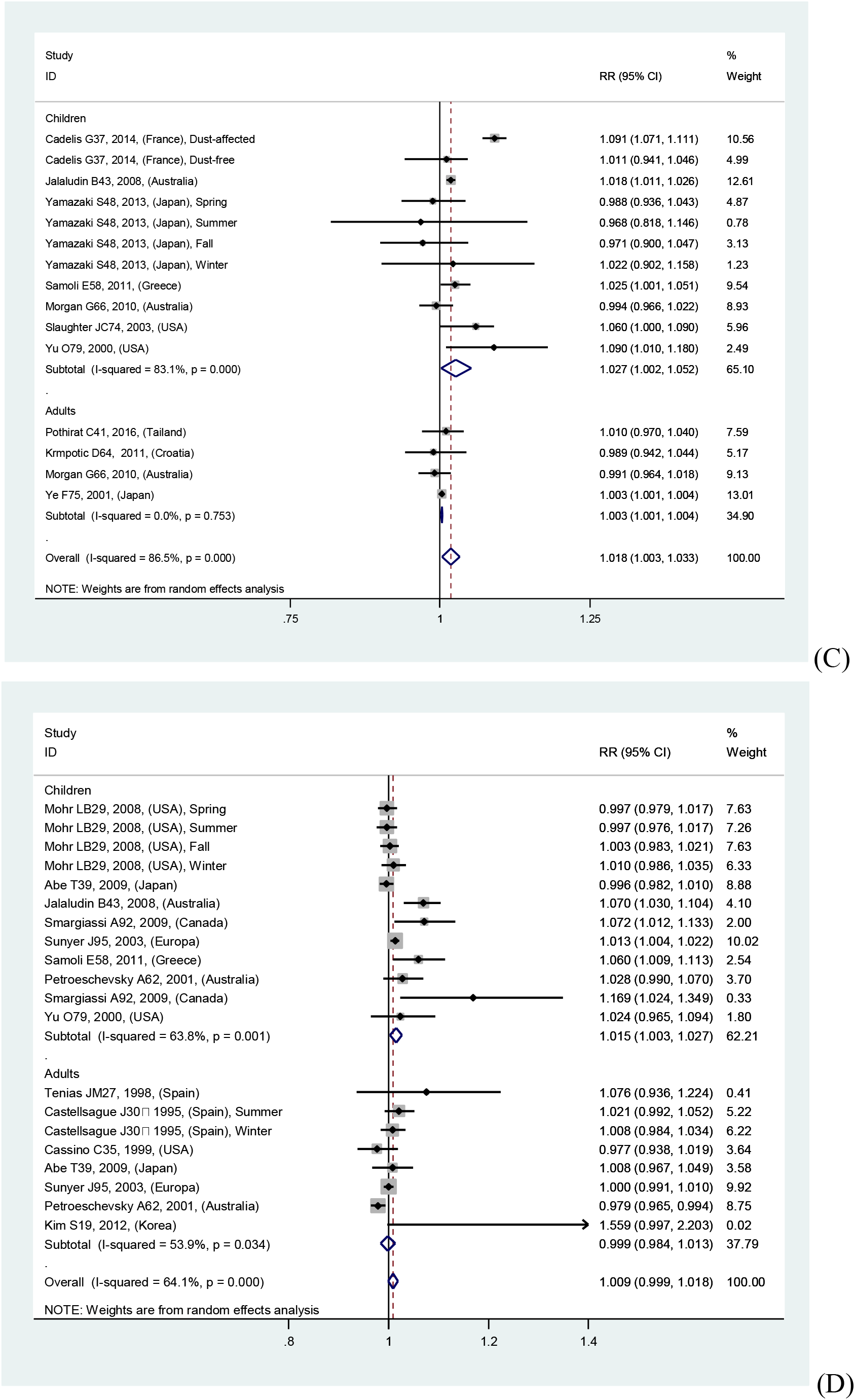
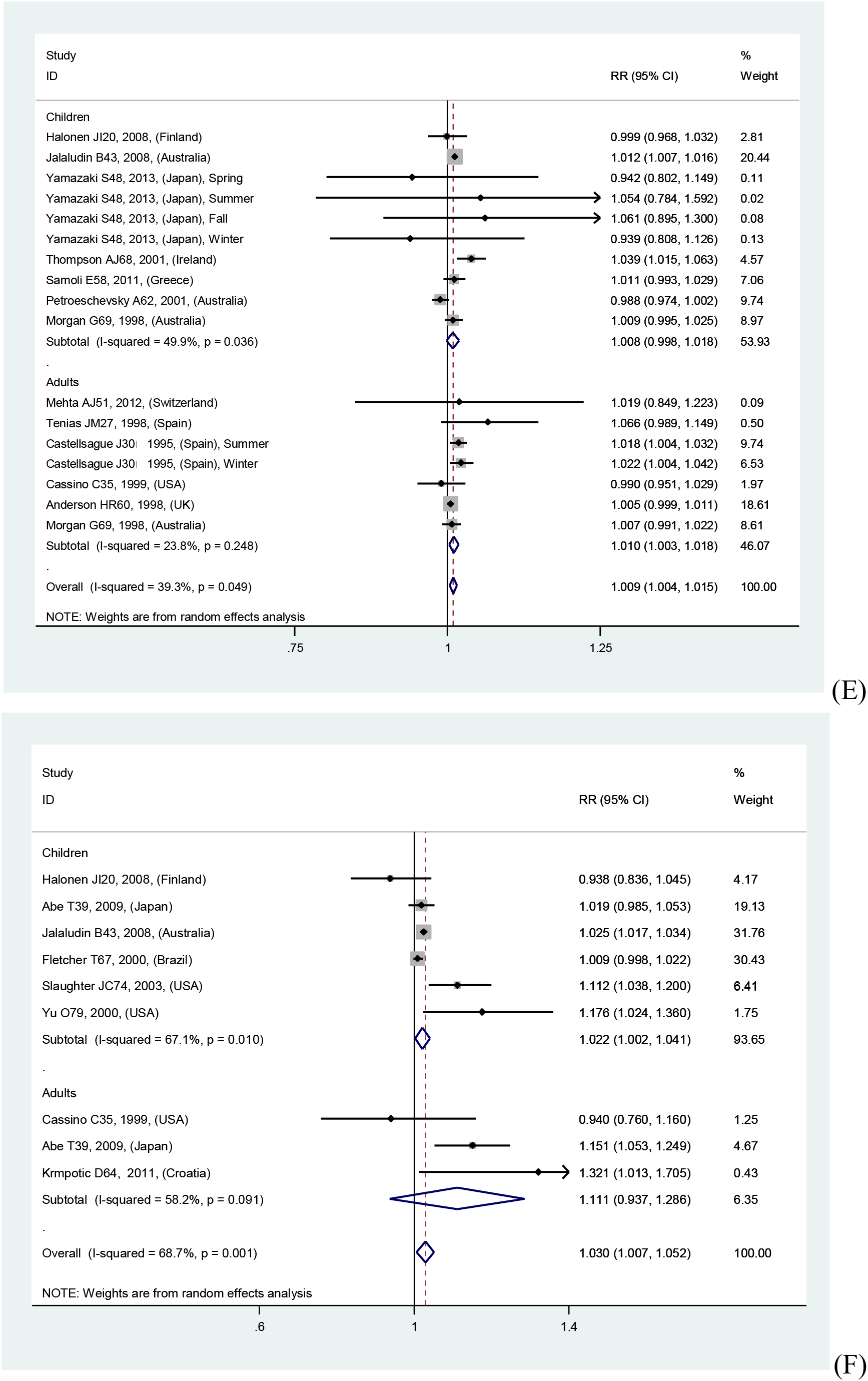
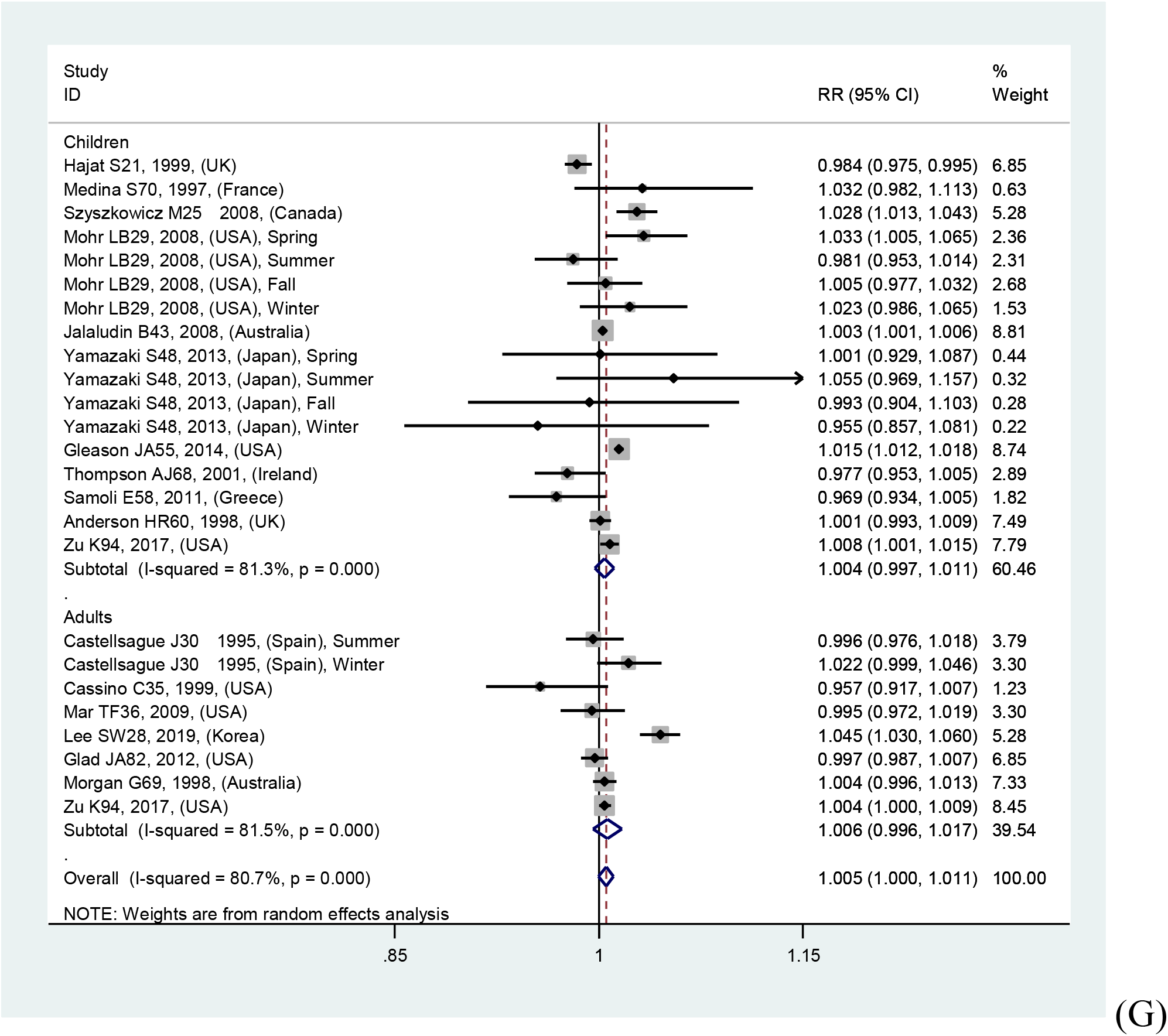

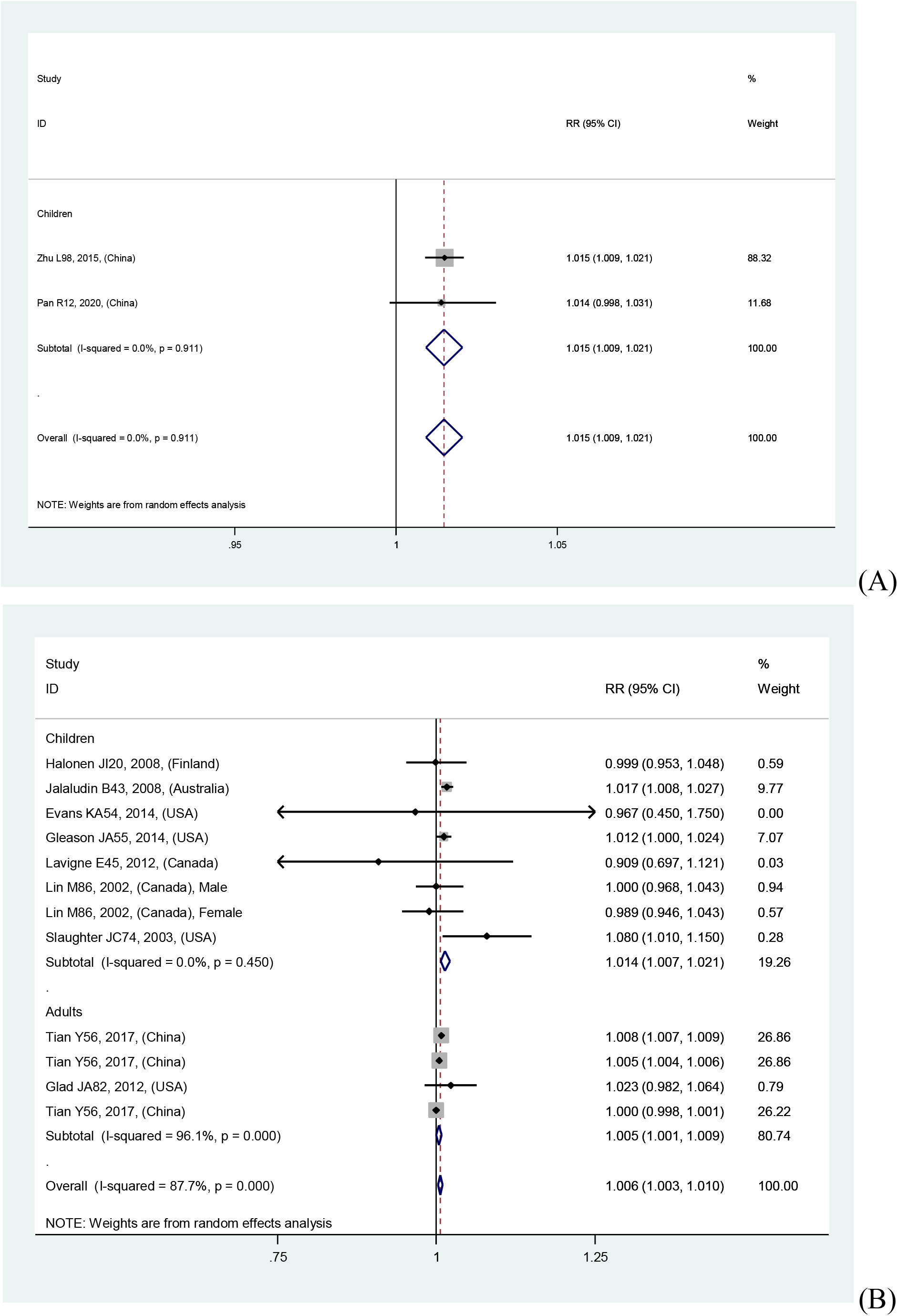
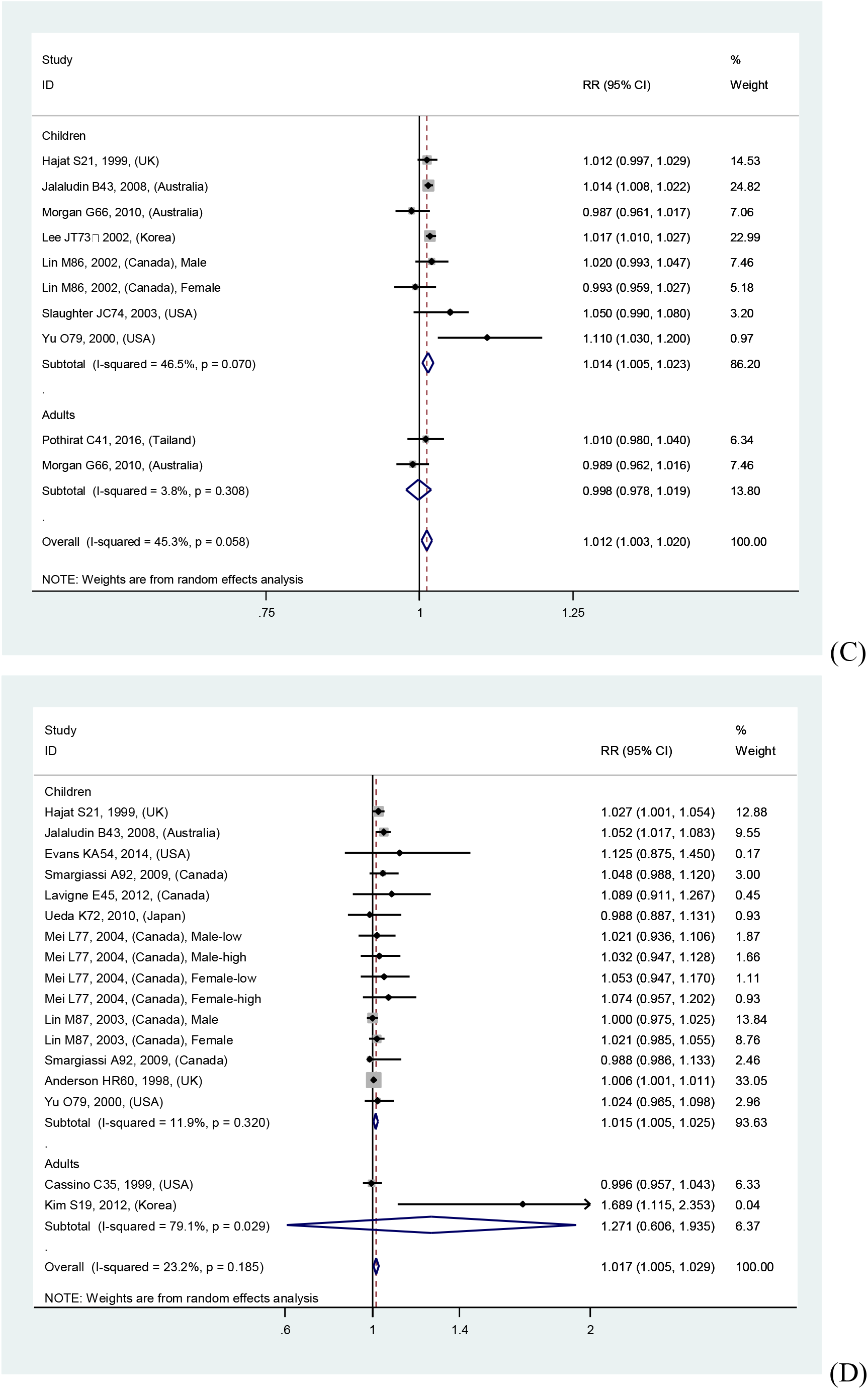
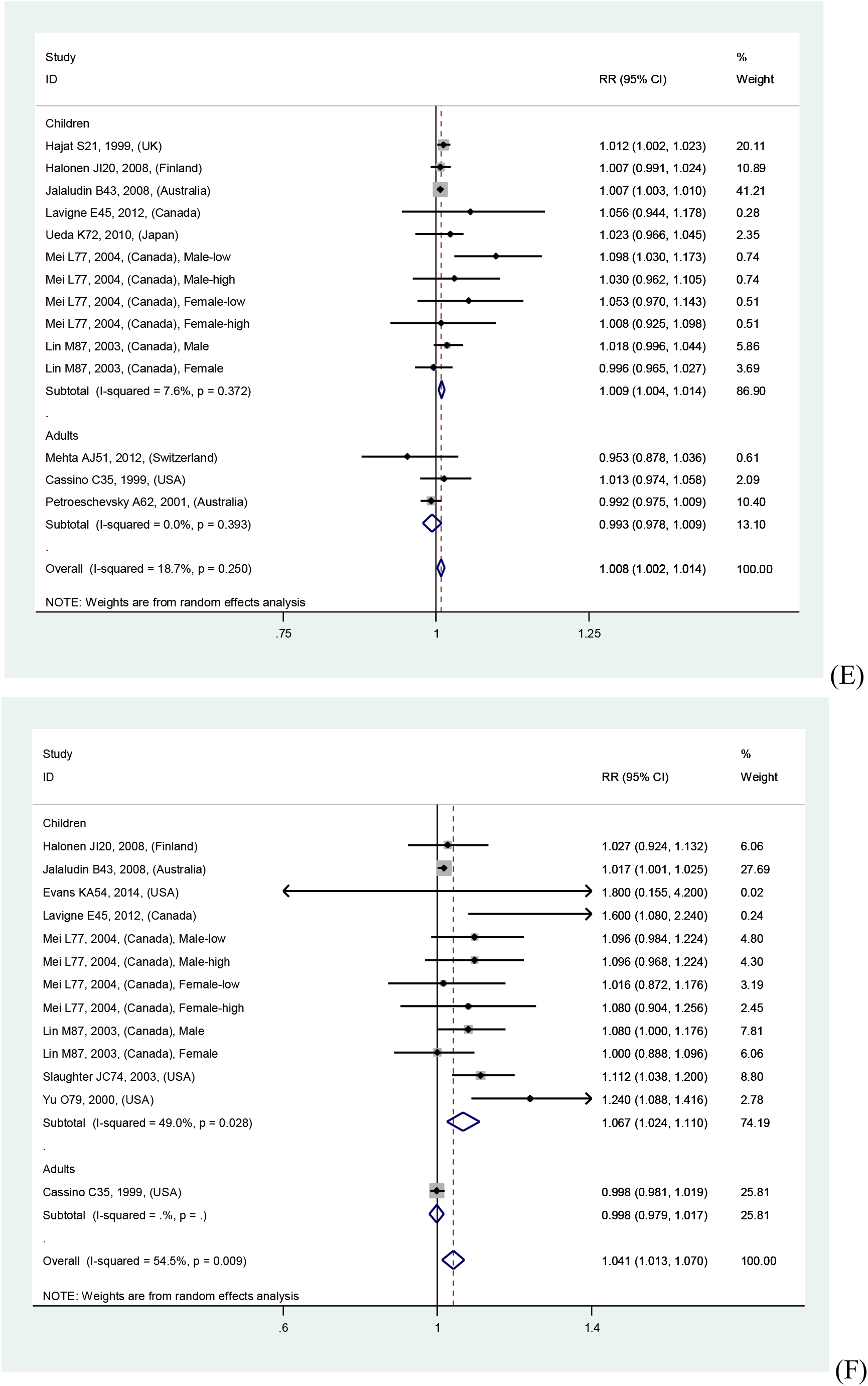
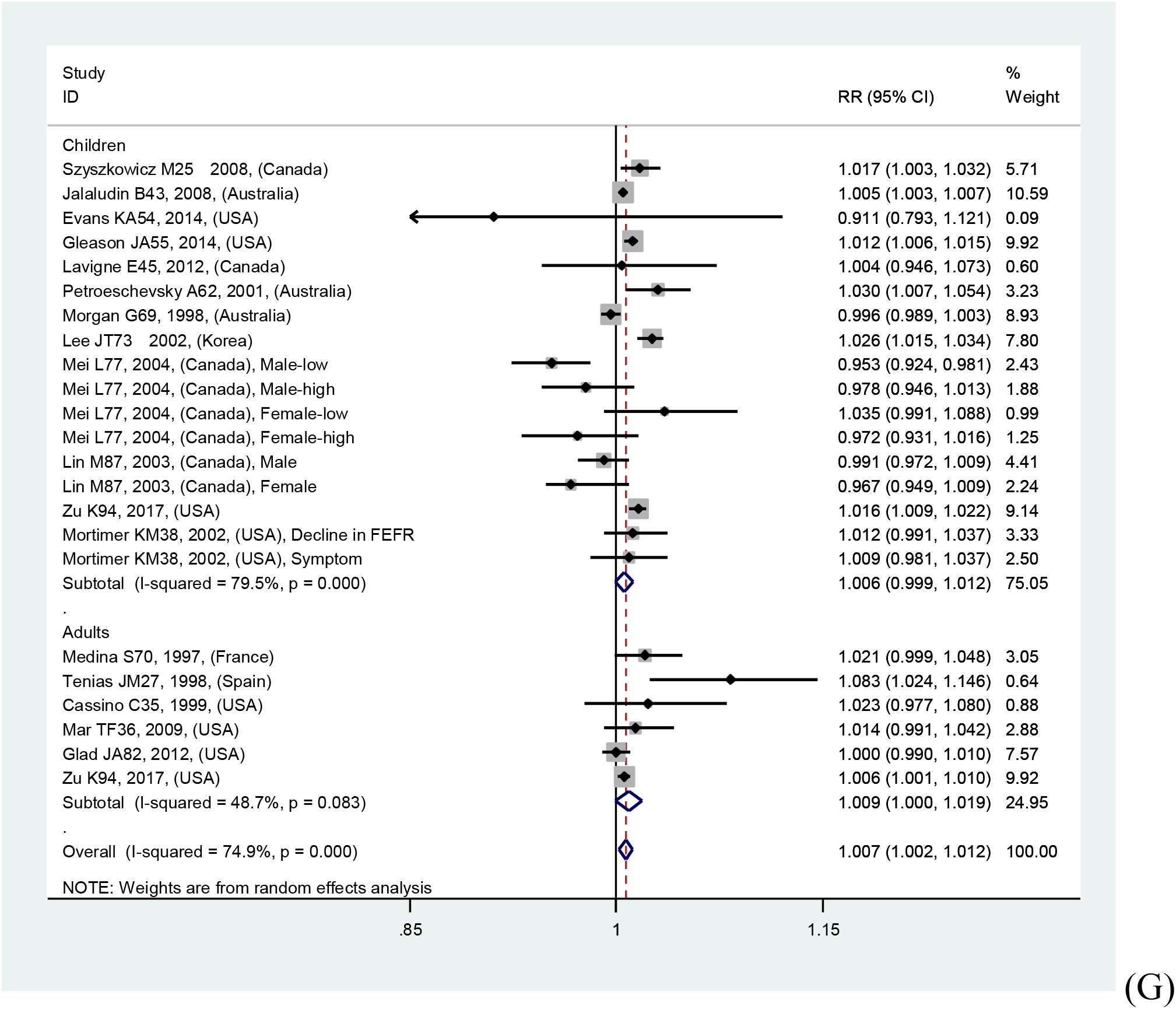

