## Supplementary material for "Outdoor air pollution and the risk of asthma exacerbations in single lag0 and lag1 exposure patterns: A systematic review and meta-analysis": Figure S1 Forest plot for relationships between air pollutants and asthma exacerbations in overall and various outcomes analyses

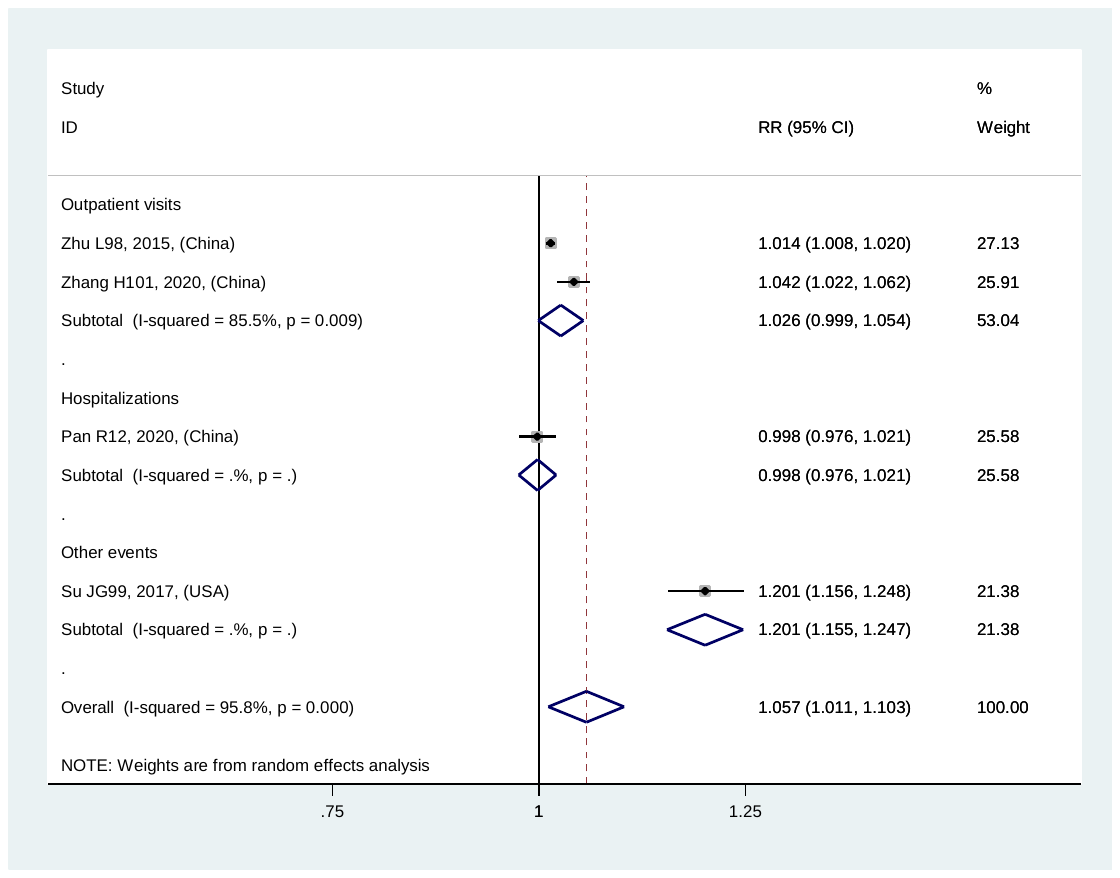
(A)

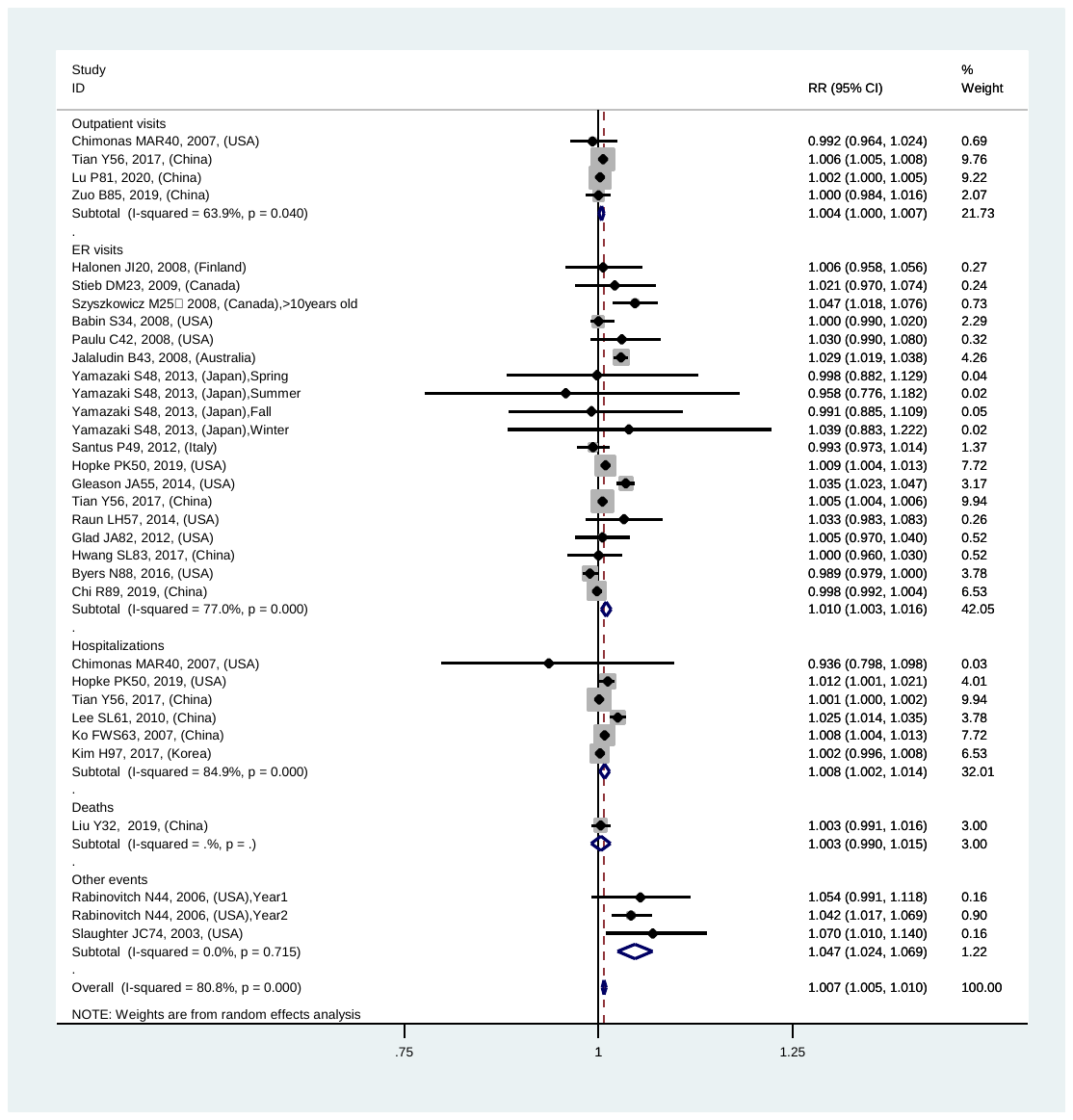
(B)

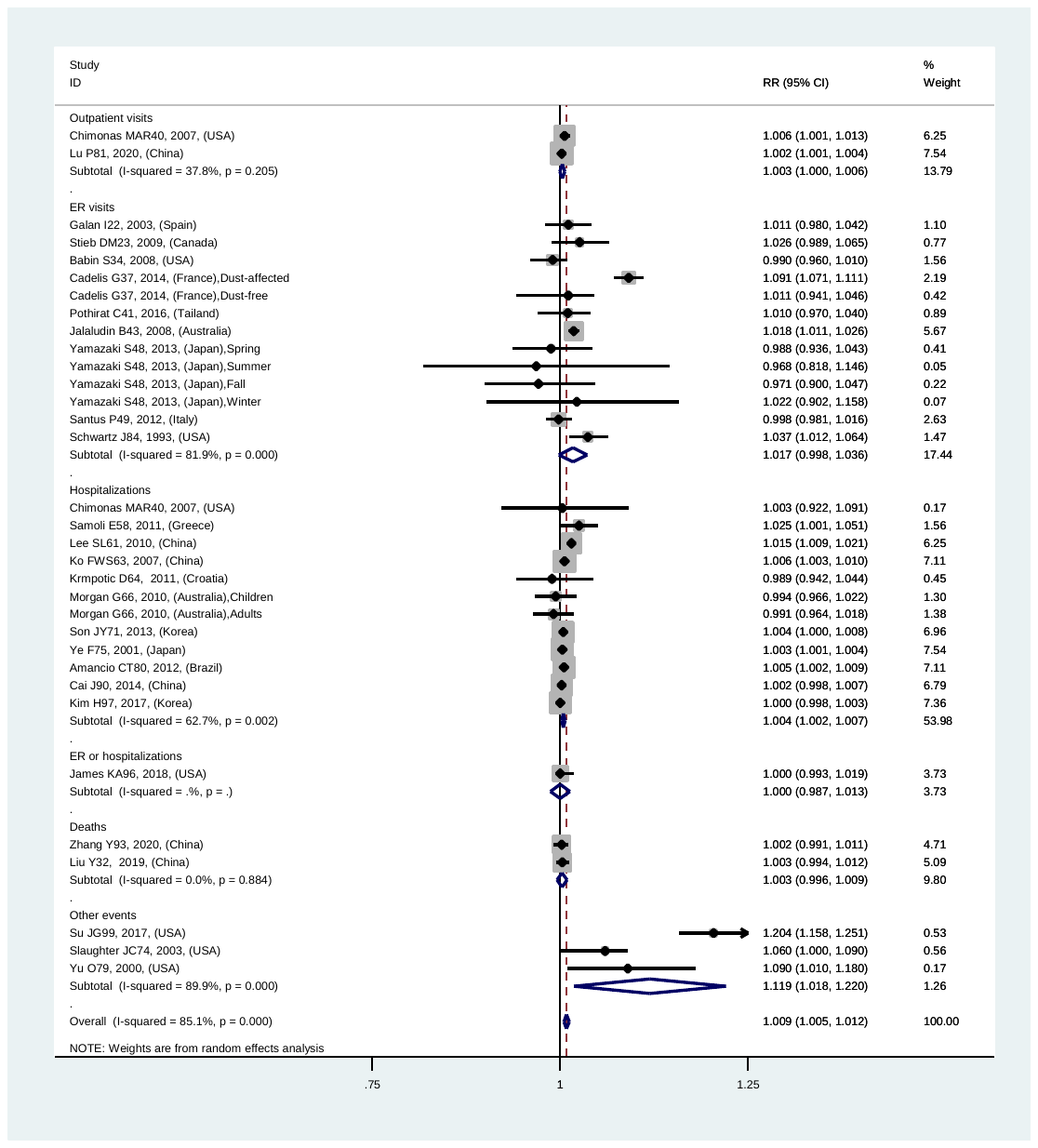
(C)

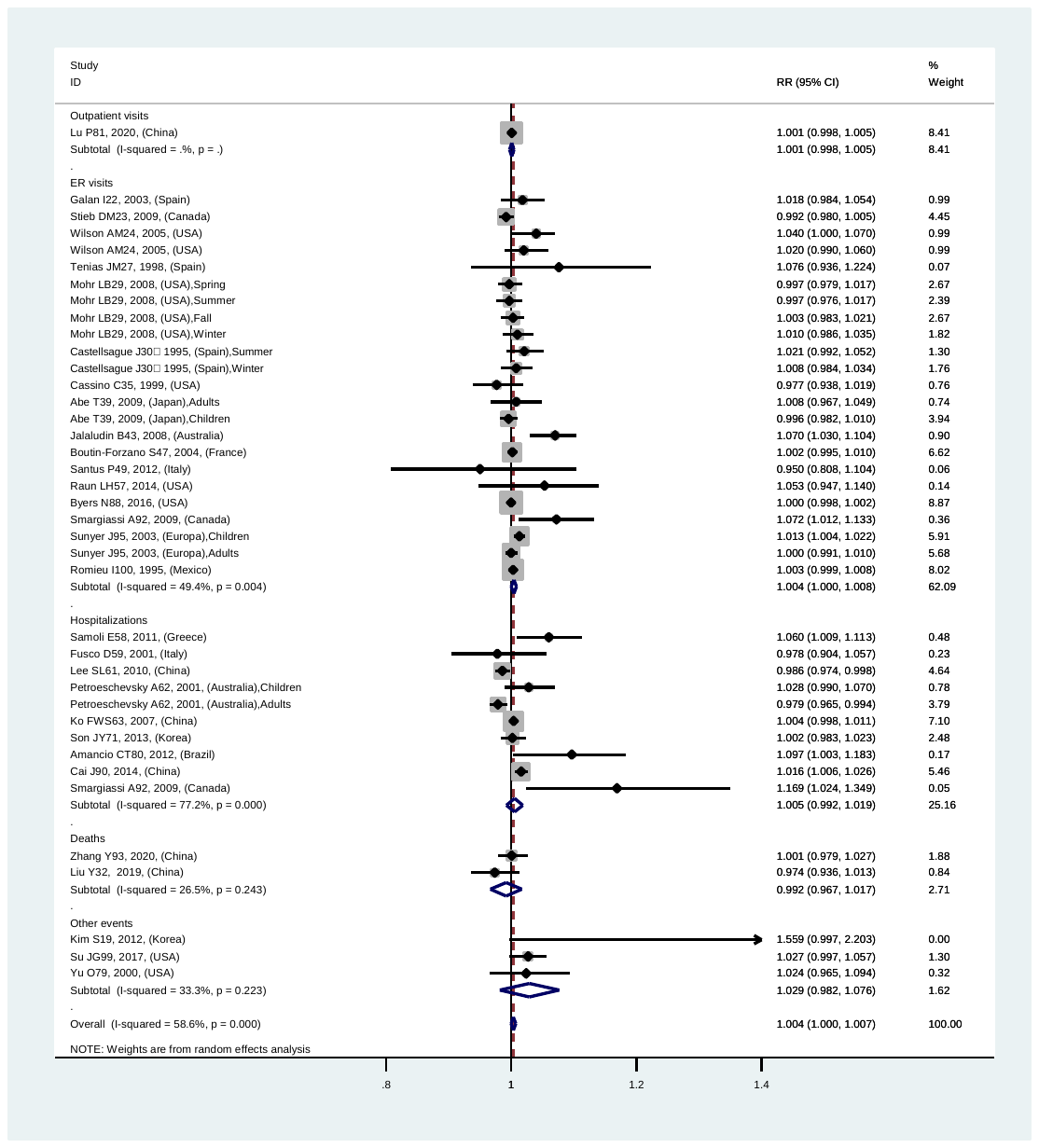
(D)

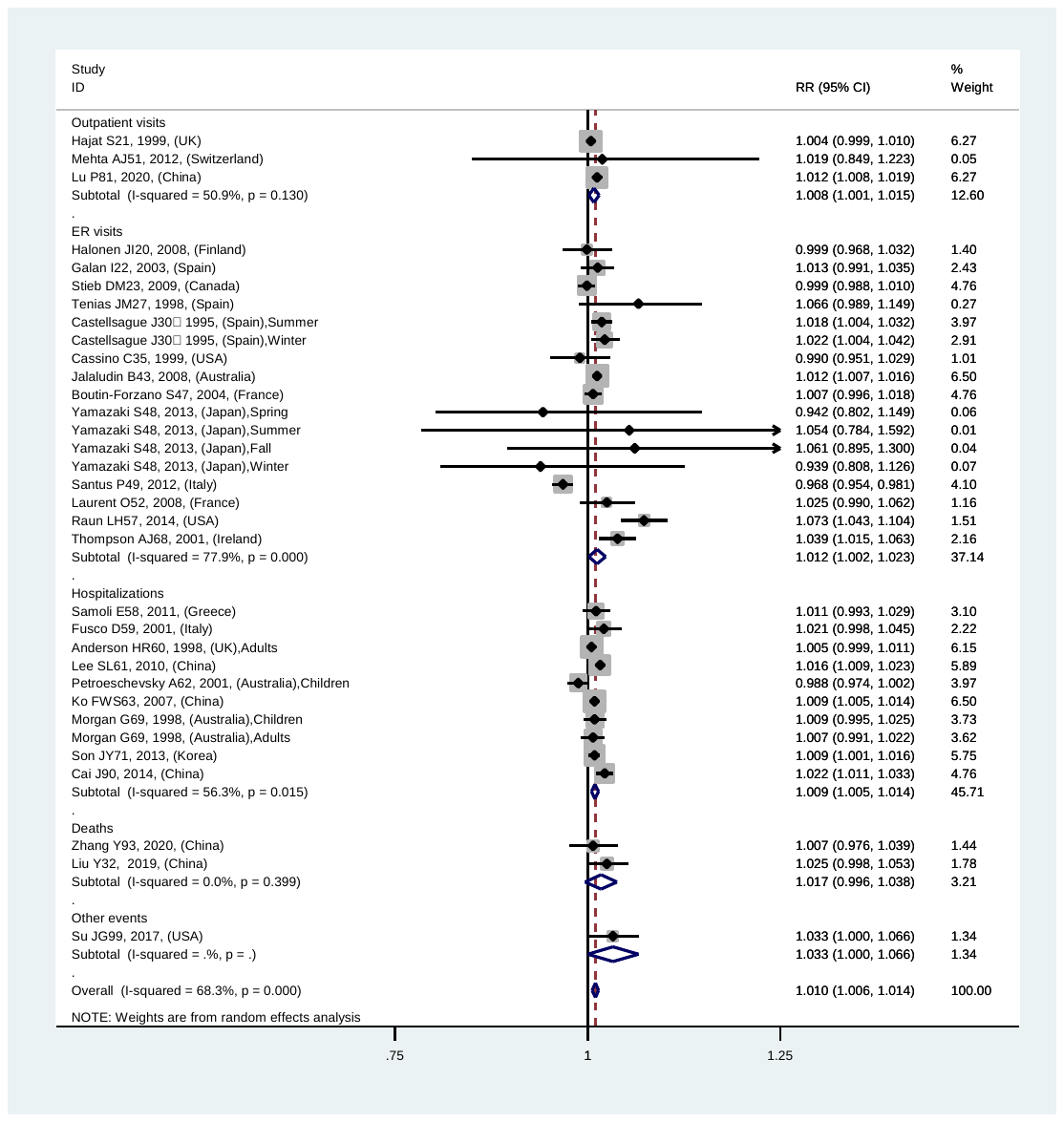
(E)

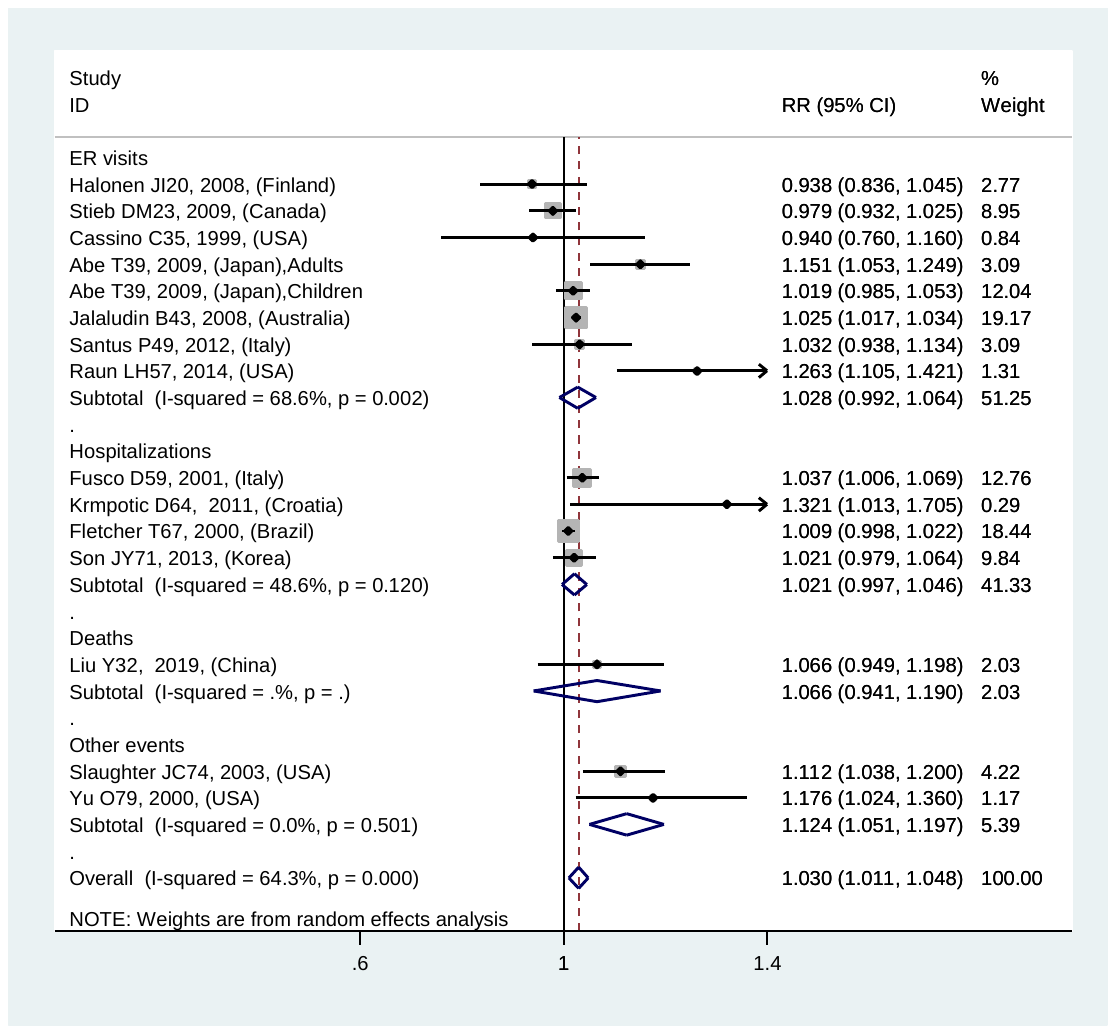
(F)

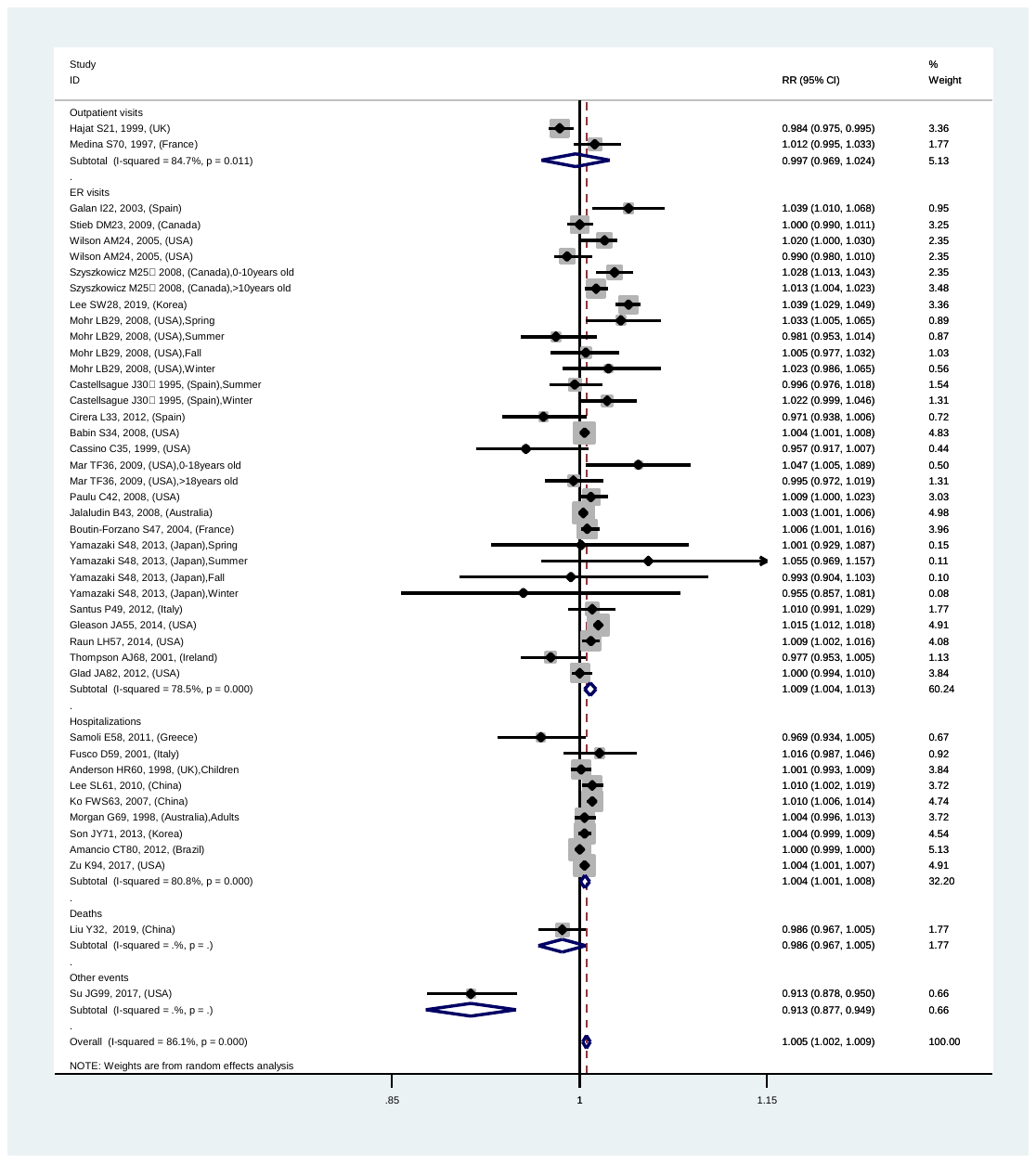
(G)

Figure S1-1 Forest plot for relationships between air pollutants ((A) AQI, (B) PM_2.5_, (C) PM_10_, (D) SO_2_, (E) NO_2_, (F) CO, (G) O_3_) and asthma exacerbations with lag0 exposure in overall and various outcomes analyses

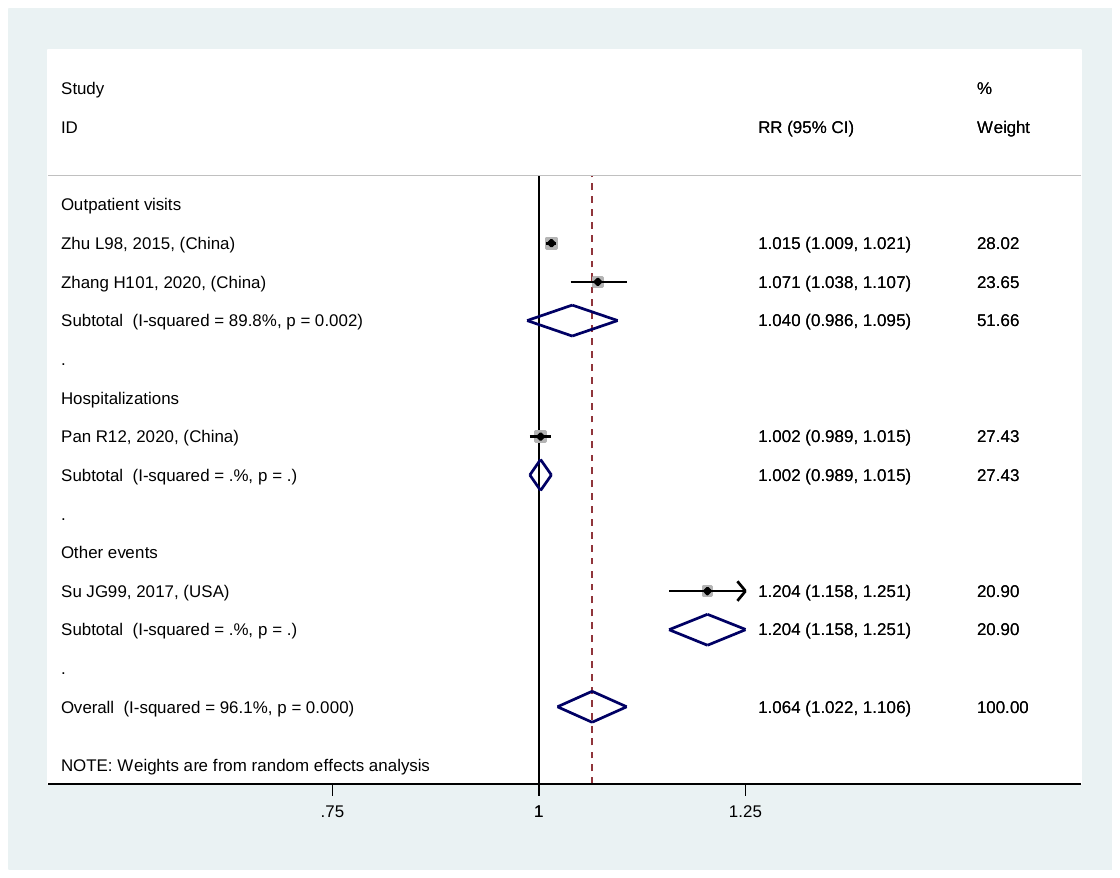
(A)

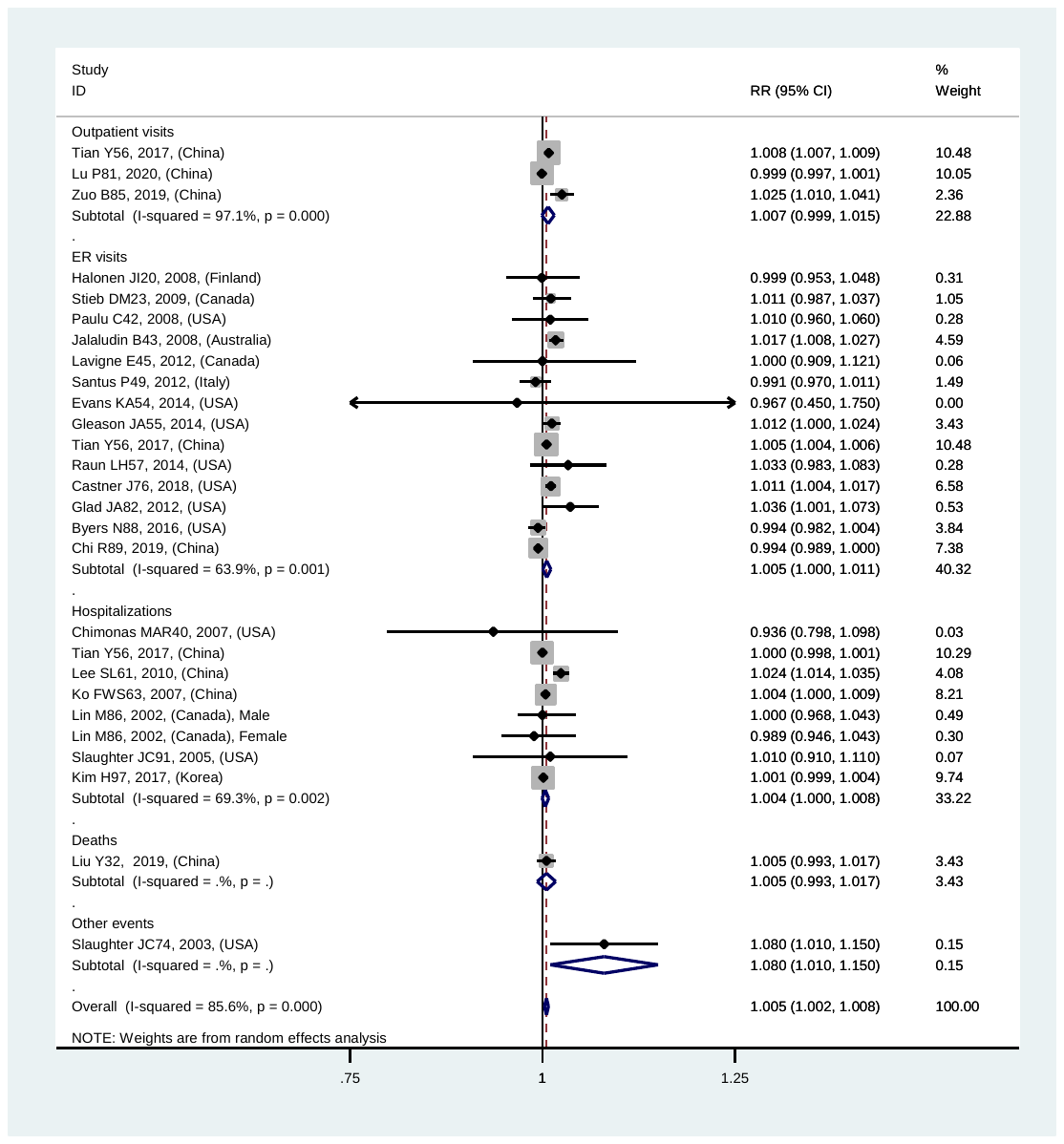
(B)

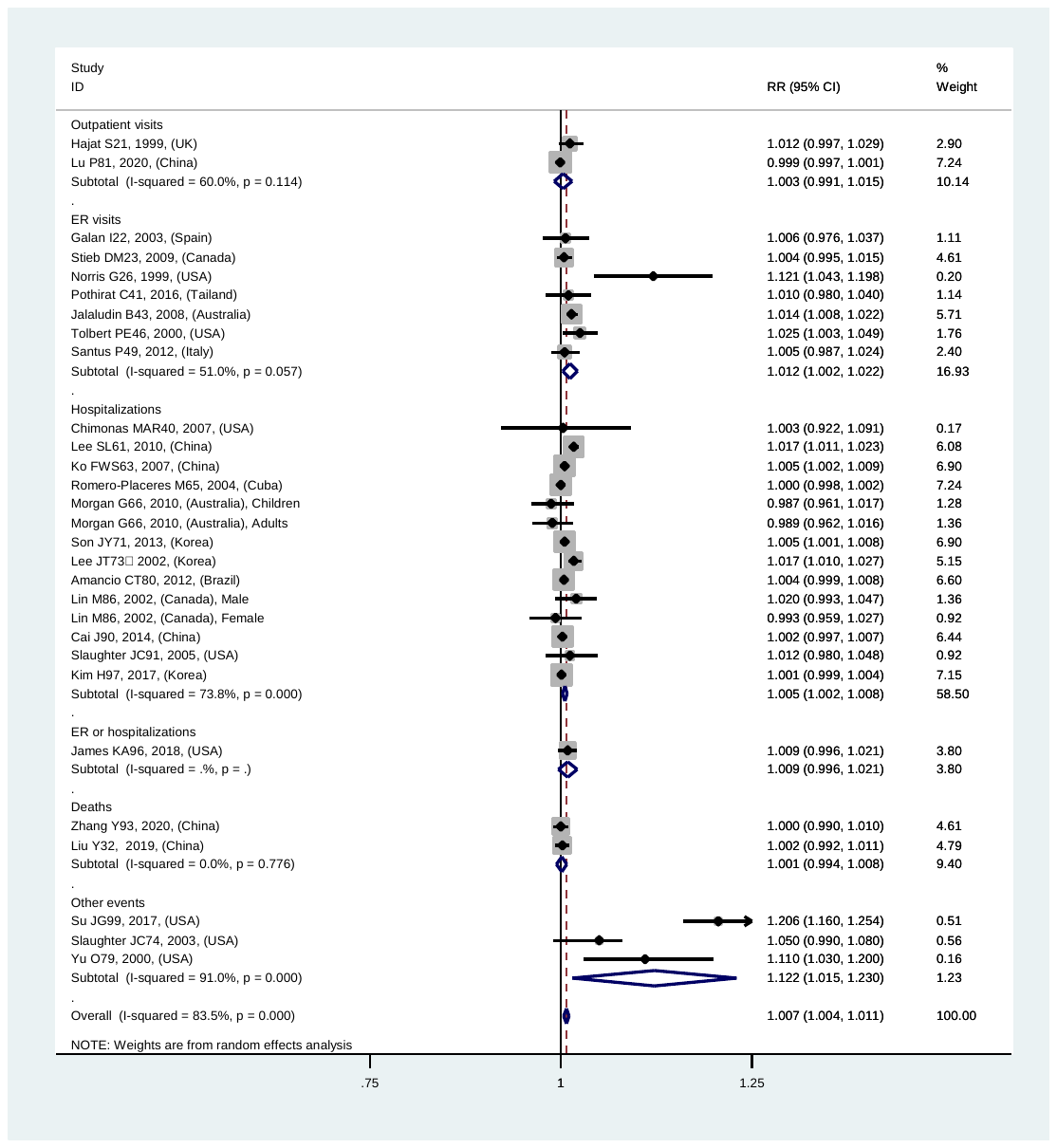
(C)

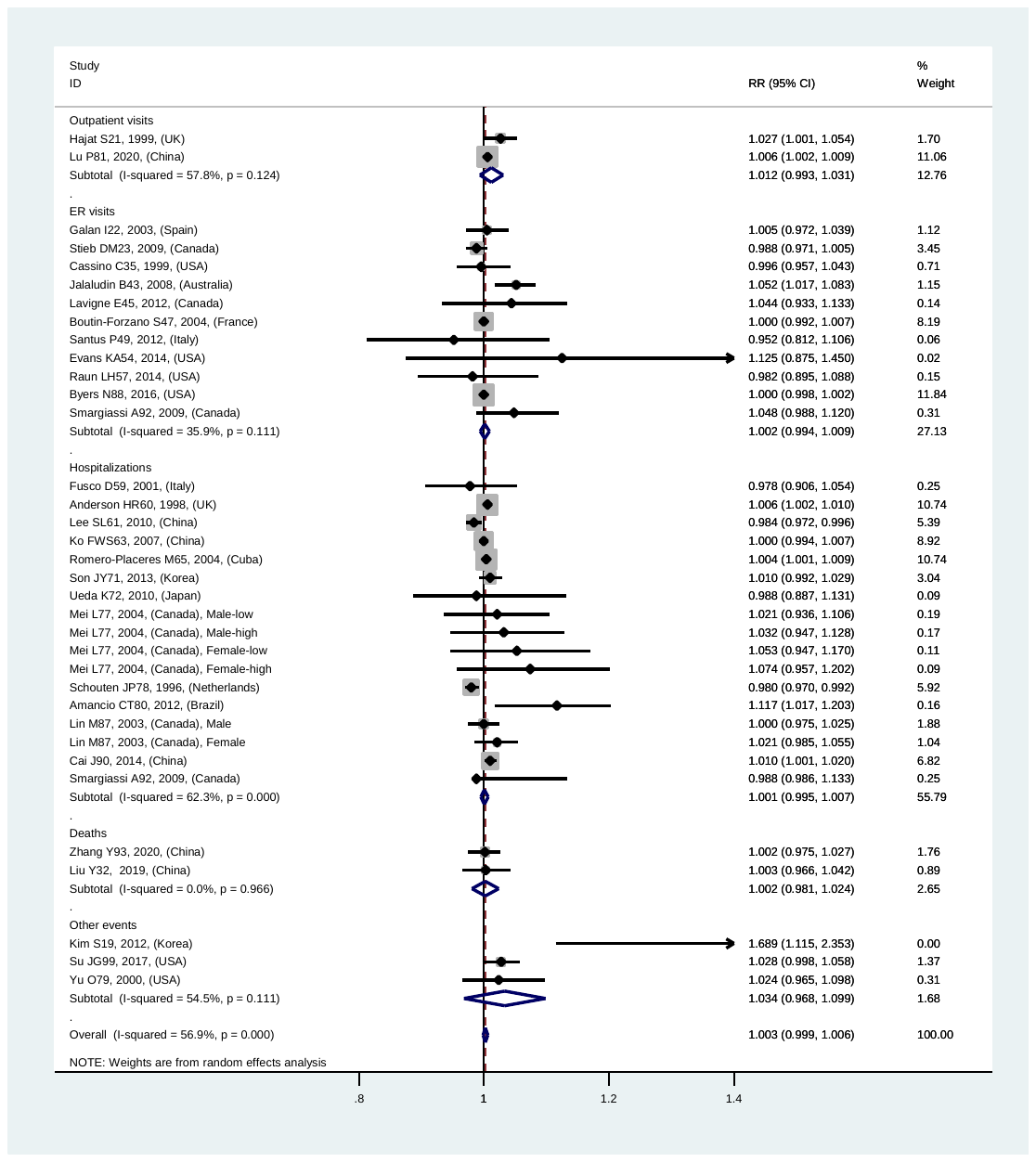
(D)

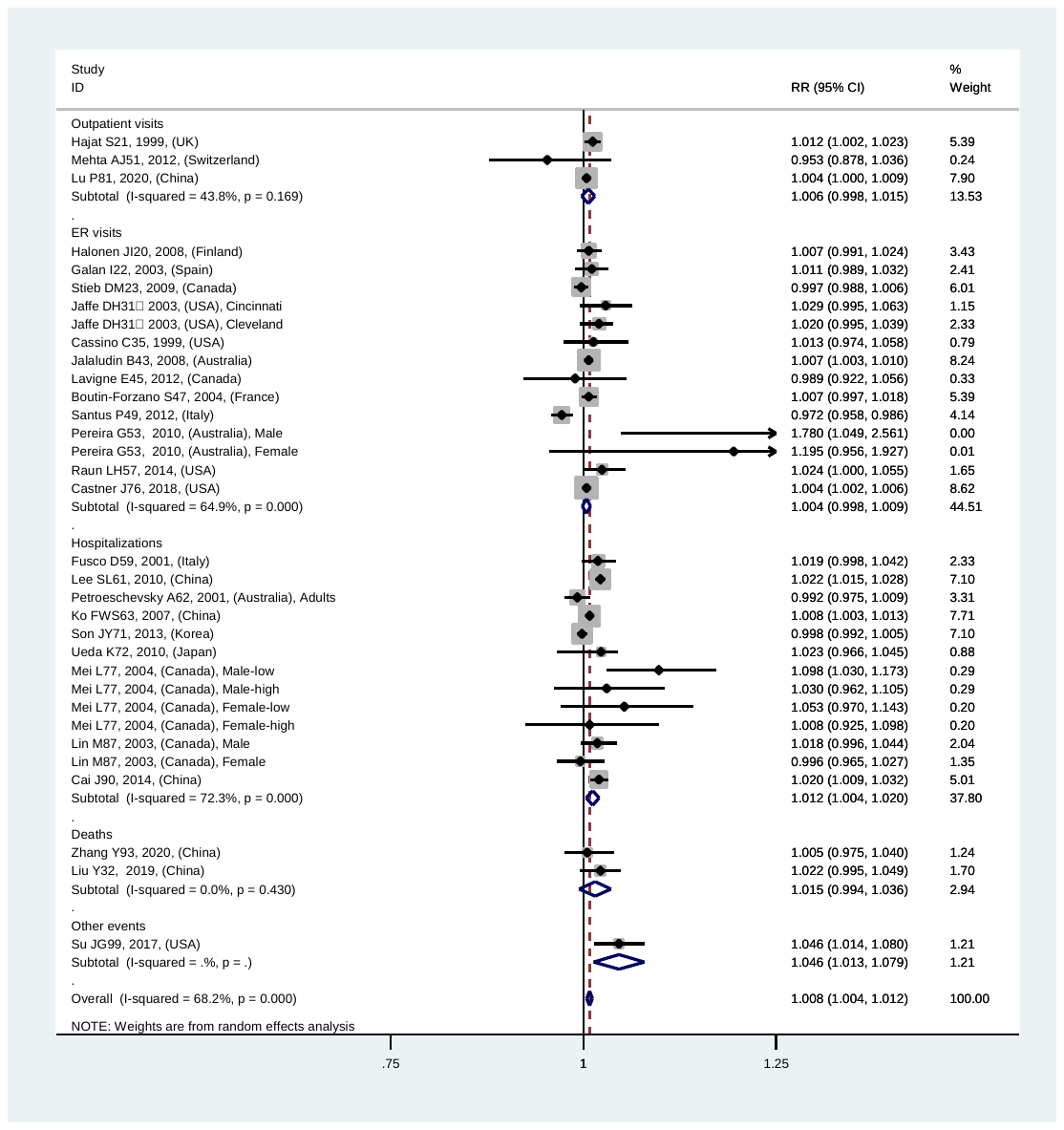
(E)

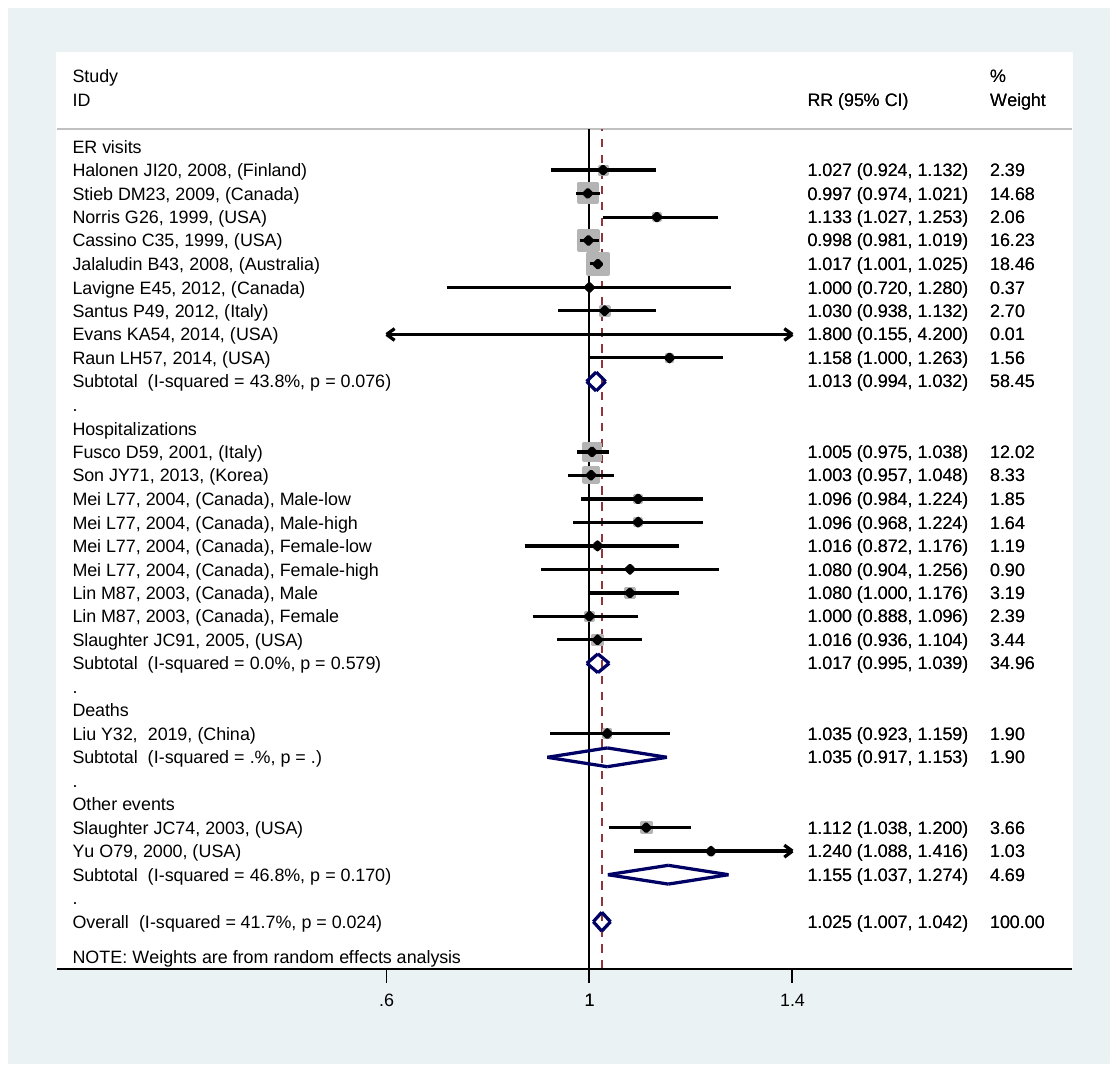
(F)

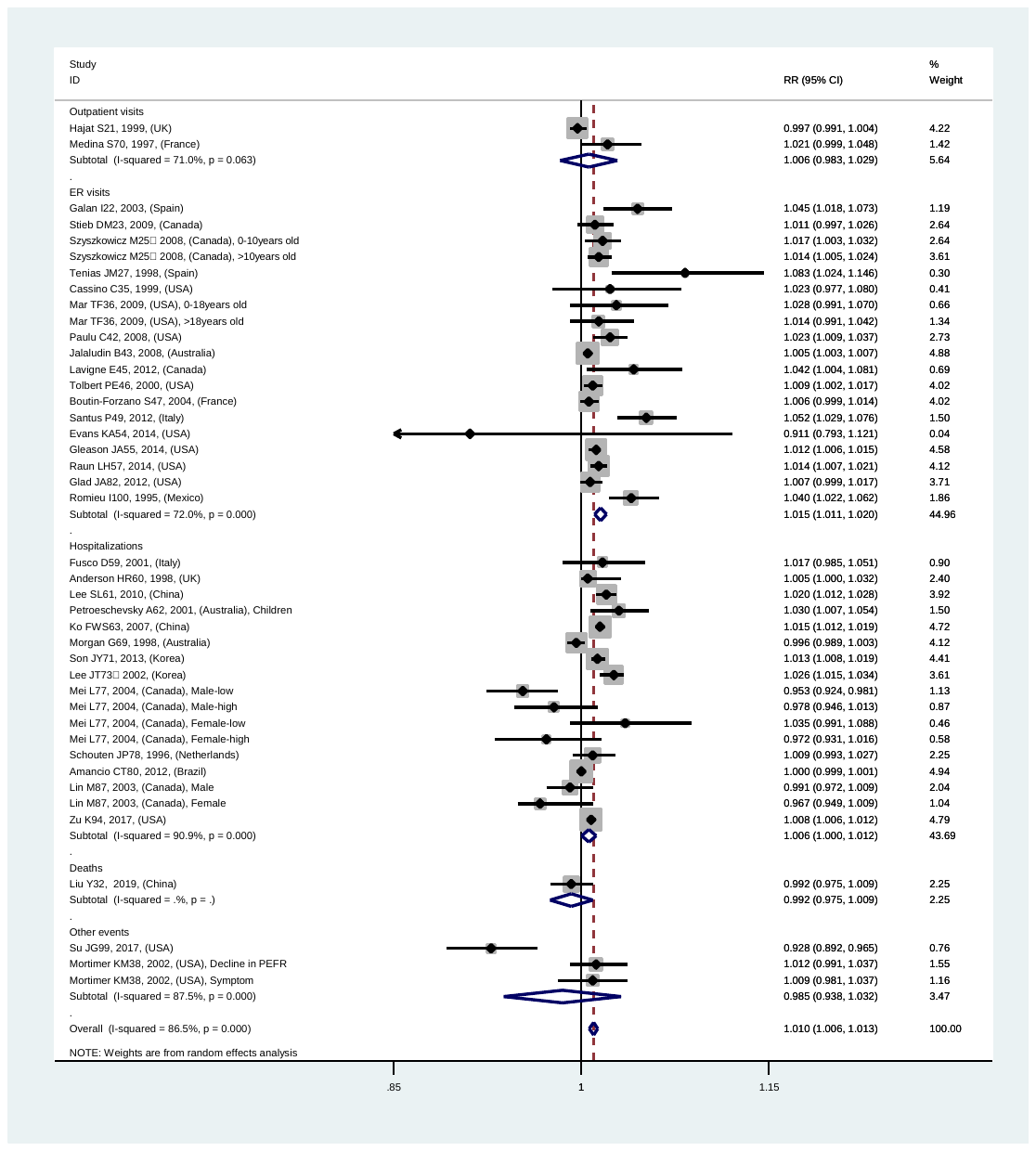
(G)

Figure S1-2 Forest plot for relationships between air pollutants ((A) AQI, (B) PM_2.5_, (C) PM_10_, (D) SO_2_, (E) NO_2_, (F) CO, (G) O_3_) and asthma exacerbations with lag1 exposure in overall and various outcomes analyses
