## Supplementary material for "Outdoor air pollution and the risk of asthma exacerbations in single lag0 and lag1 exposure patterns: A systematic review and meta-analysis": Figure S2. Begg's funnel plot for relationships between air pollutants and asthma exacerbations in overall analyses

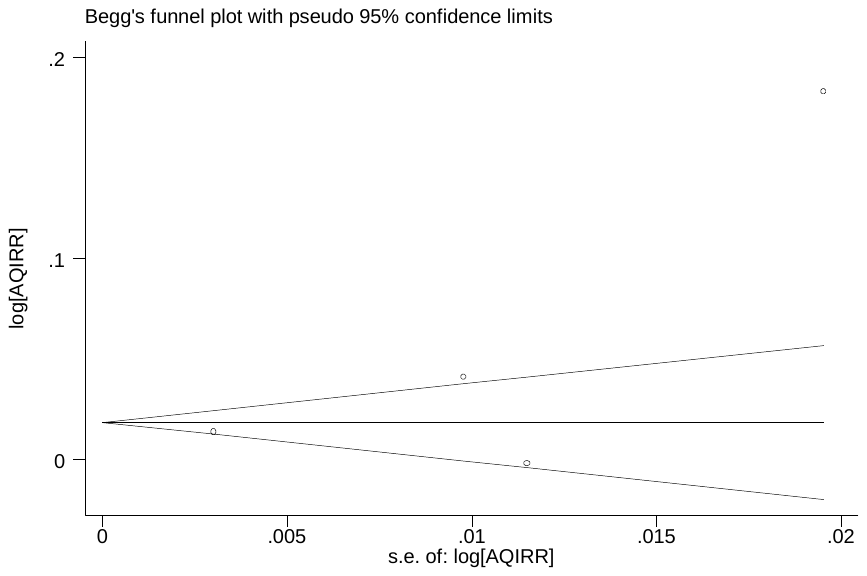

Figure S2-1 Begg’s funnel plot for relationship between AQI and asthma exacerbations with lag0 exposure in overall analyses (p=0.308)

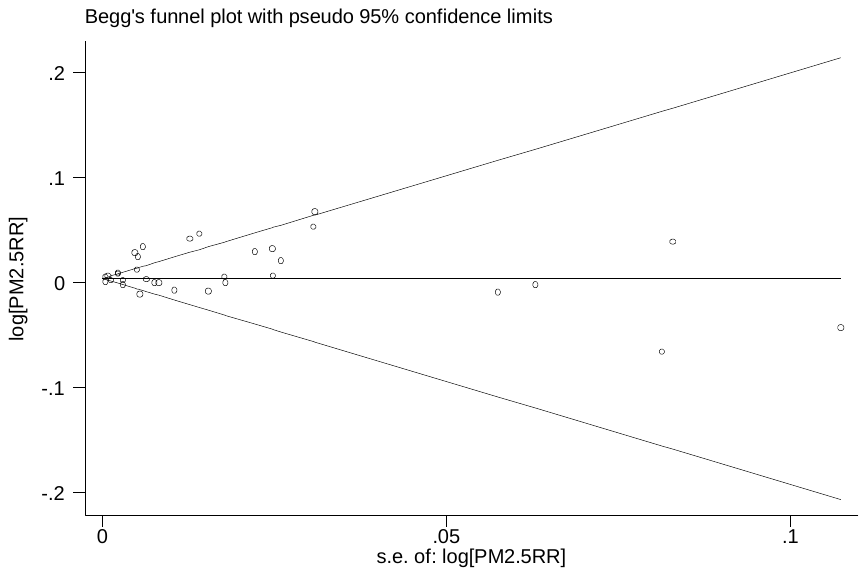

Figure S2-2 Begg’s funnel plot for relationship between PM_2.5_ and asthma exacerbations with lag0 exposure in overall analyses (p=0.687)

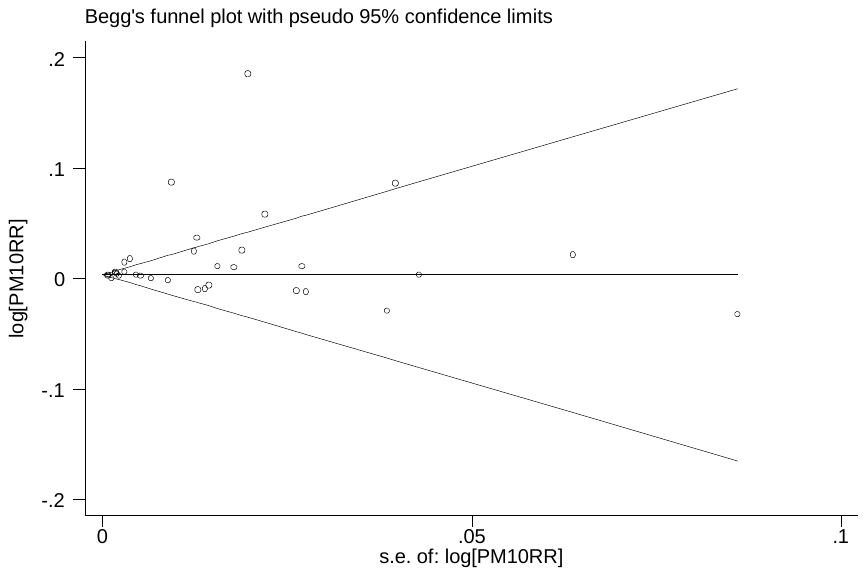

Figure S2-3 Begg’s funnel plot for relationship between PM_10_ and asthma exacerbations with lag0 exposure in overall analyses (p=0.664)

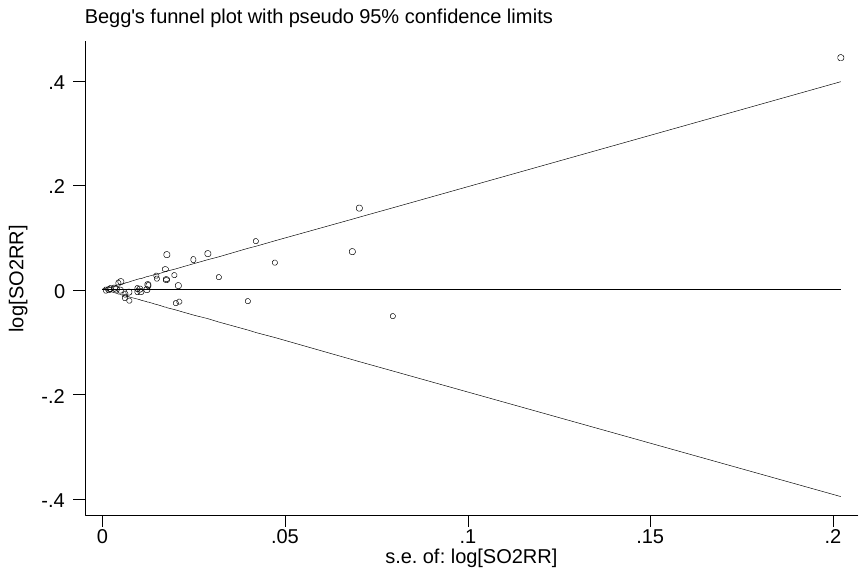

Figure S2-4 Begg’s funnel plot for relationship between SO_2_ and asthma exacerbations with lag0 exposure in overall analyses (p=0.026)

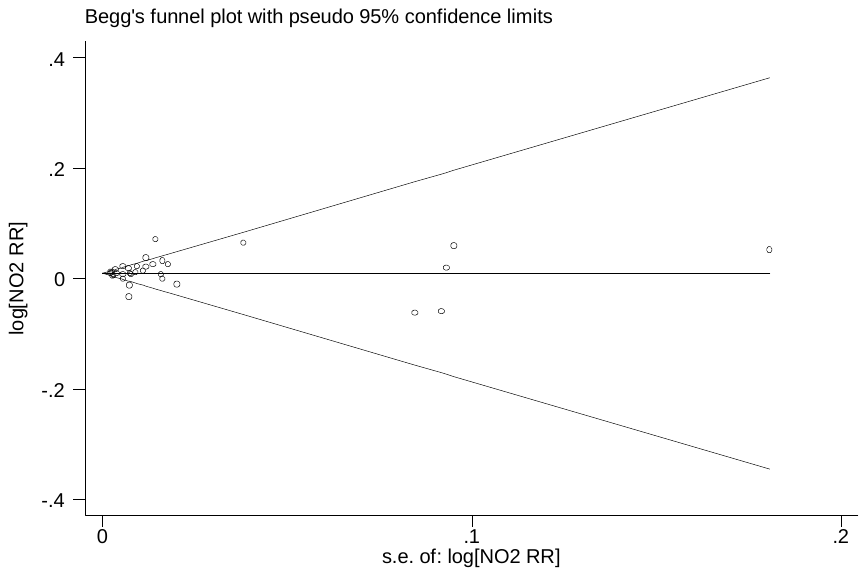

Figure S2-5 Begg’s funnel plot for relationship between NO_2_ and asthma exacerbations with lag0 exposure in overall analyses (p=0.631)

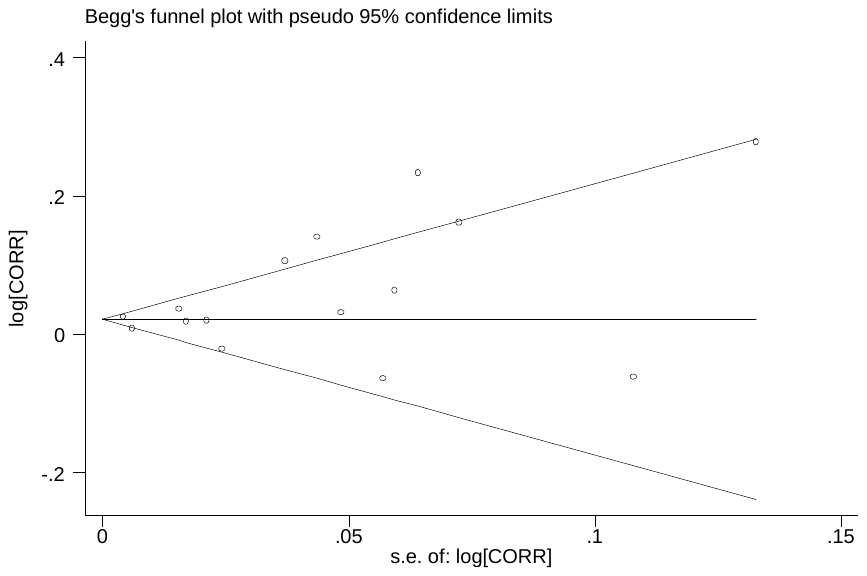

Figure S2-6 Begg’s funnel plot for relationship between CO and asthma exacerbations with lag0 exposure in overall analyses (p=0.322)

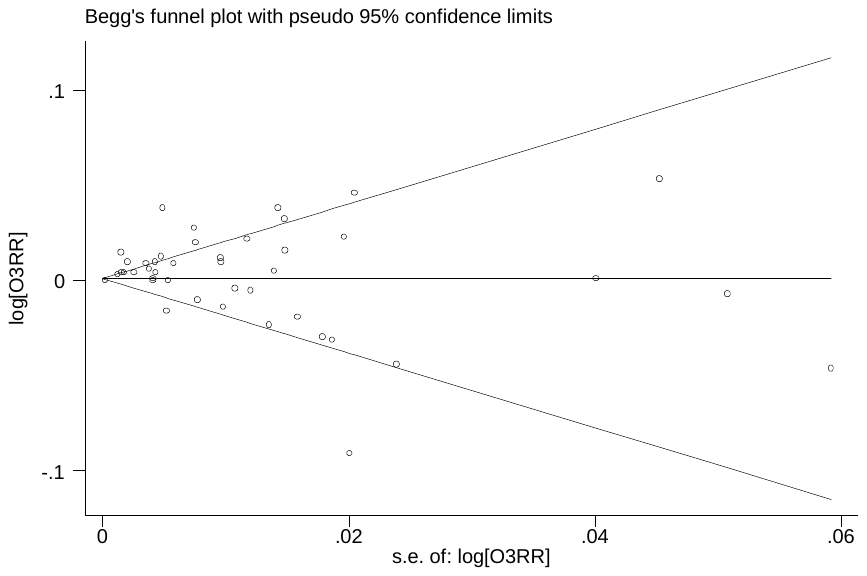

Figure S2-7 Begg’s funnel plot for relationship between O_3_ and asthma exacerbations with lag0 exposure in overall analyses (p=0.015)

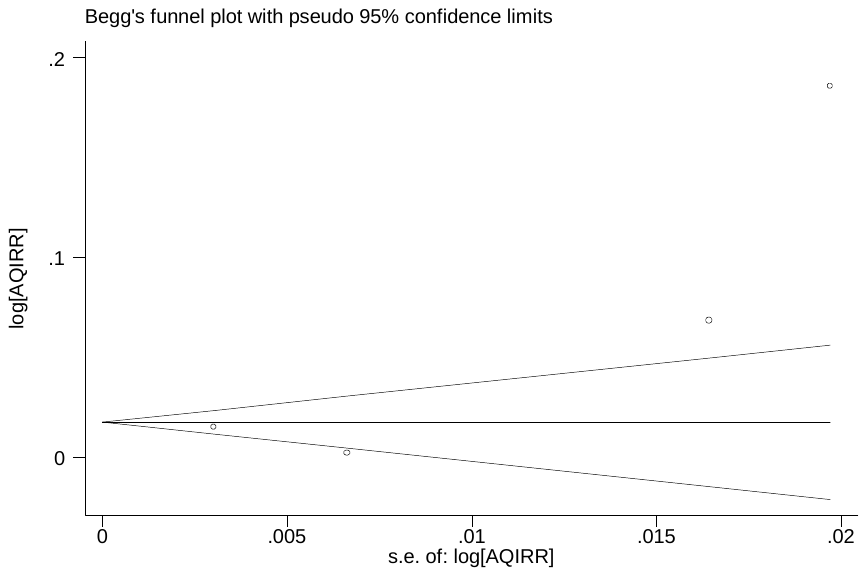

Figure S2-8 Begg’s funnel plot for relationship between AQI and asthma exacerbations with lag1 exposure in overall analyses (p=0.308)

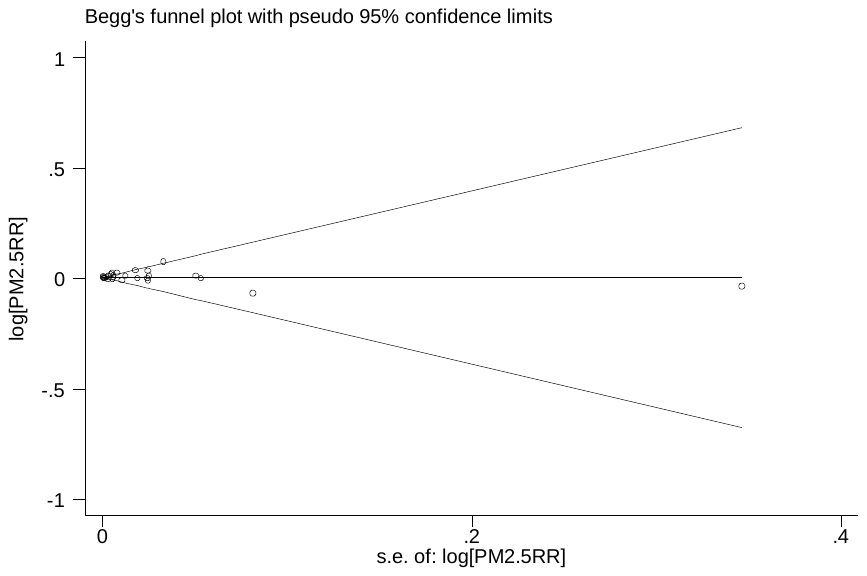

Figure S2-9 Begg’s funnel plot for relationship between PM_2.5_ and asthma exacerbations with lag1 exposure in overall analyses (p=0.835)

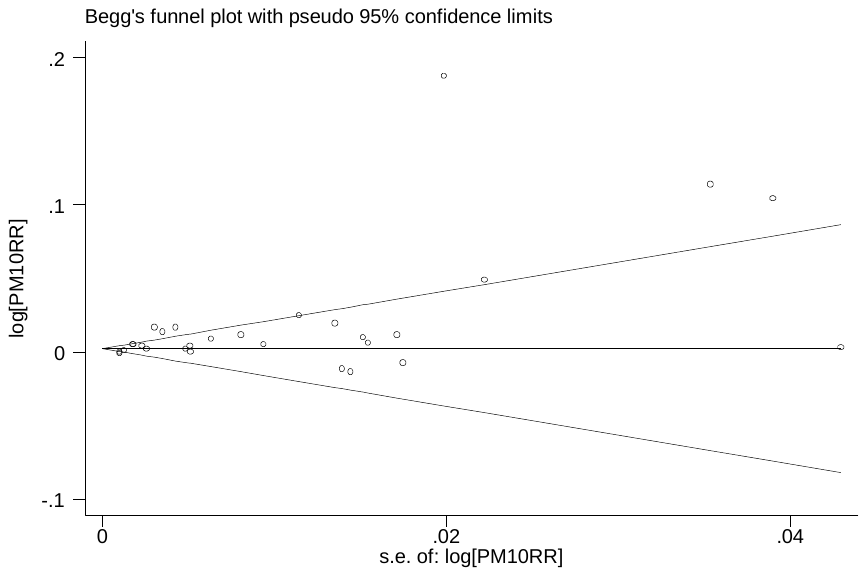

Figure S2-10 Begg’s funnel plot for relationship between PM_10_ and asthma exacerbations with lag1 exposure in overall analyses (p=0.253)

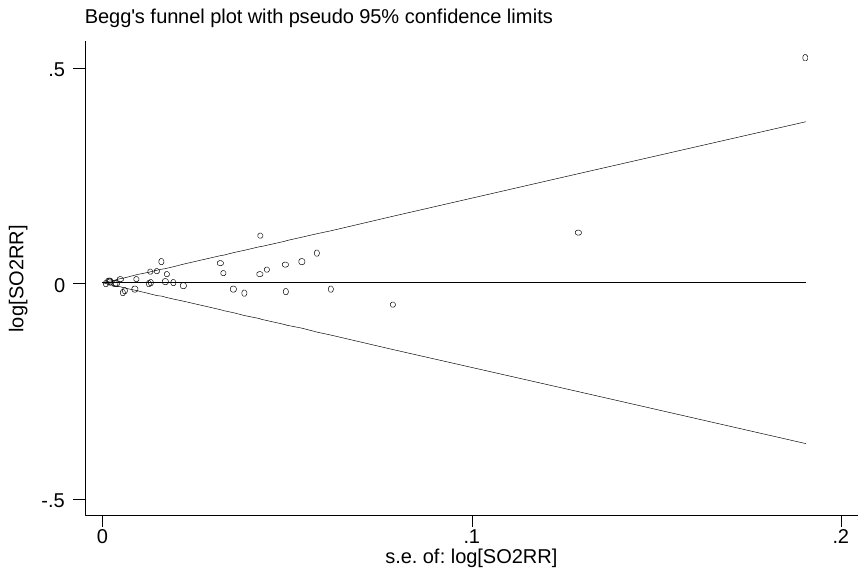

Figure S2-11 Begg’s funnel plot for relationship between SO_2_ and asthma exacerbations with lag1 exposure in overall analyses (p=0.478)

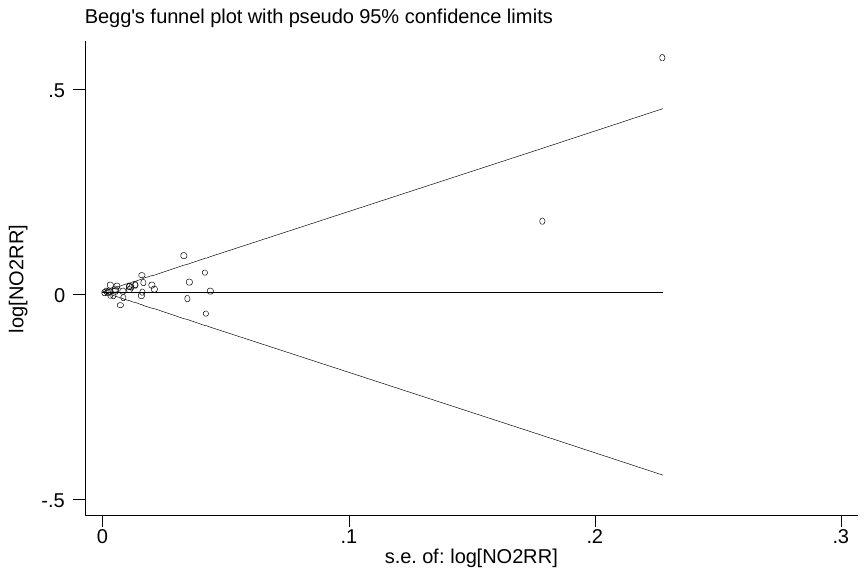

Figure S2-12 Begg’s funnel plot for relationship between NO_2_ and asthma exacerbations with lag1 exposure in overall analyses (p=0.168)

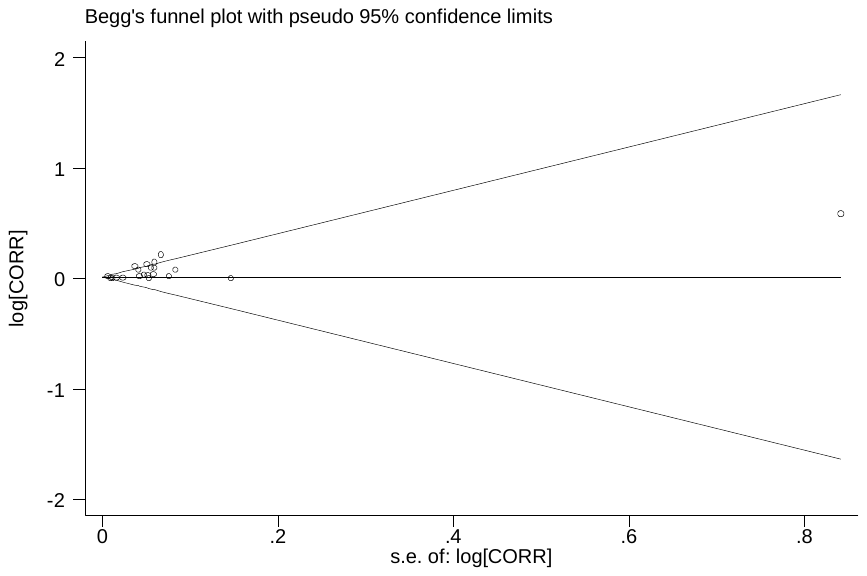

Figure S2-13 Begg’s funnel plot for relationship between CO and asthma exacerbations with lag1 exposure in overall analyses (p=0.124)

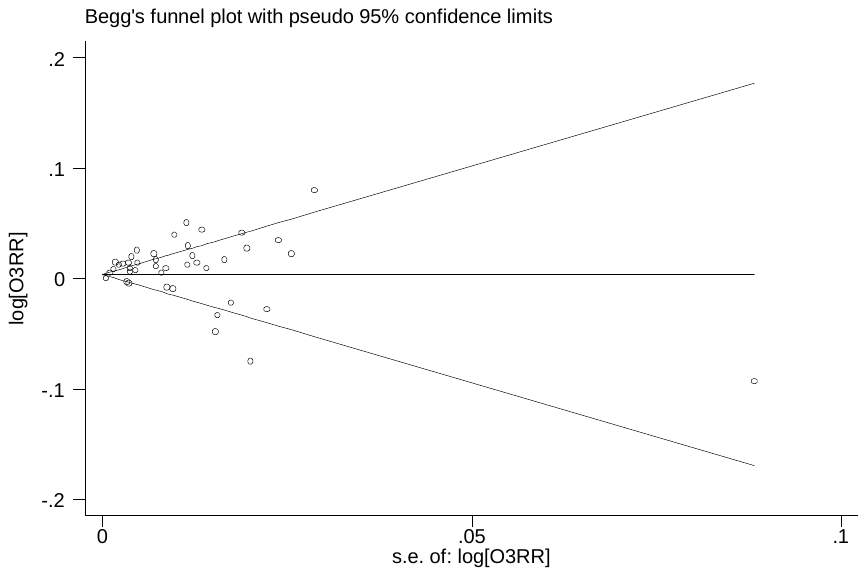

Figure S2-14 Begg’s funnel plot for relationship between O_3_ and asthma exacerbations with lag1 exposure in overall analyses (p=0.059)
