## Supplementary material for "Outdoor air pollution and the risk of asthma exacerbations in single lag0 and lag1 exposure patterns: A systematic review and meta-analysis": Figure S4. Begg's funnel plot for relationships between air pollutants and asthma exacerbations in quality sensitivity analyses

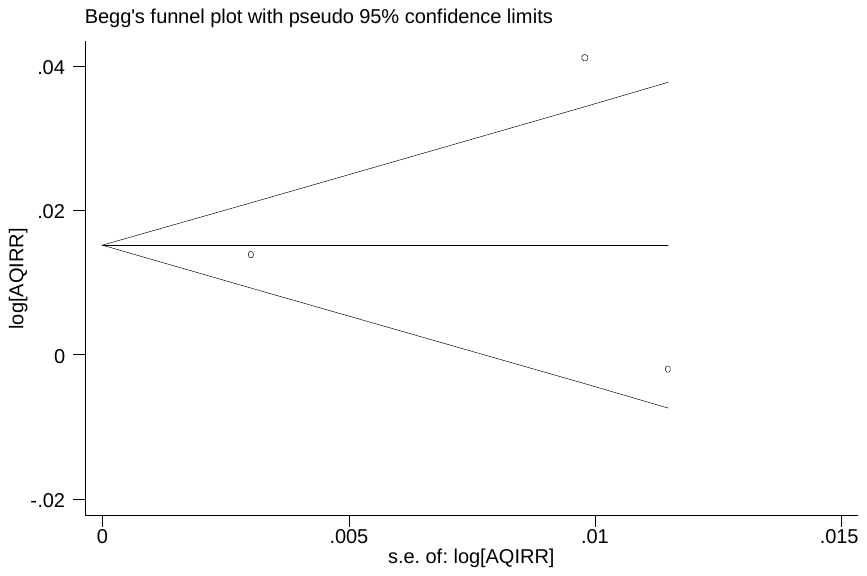

Figure S4-1 Begg’s funnel plot for relationship between AQI and asthma exacerbations with lag0 exposure in quality sensitivity analyses (p=1.000)

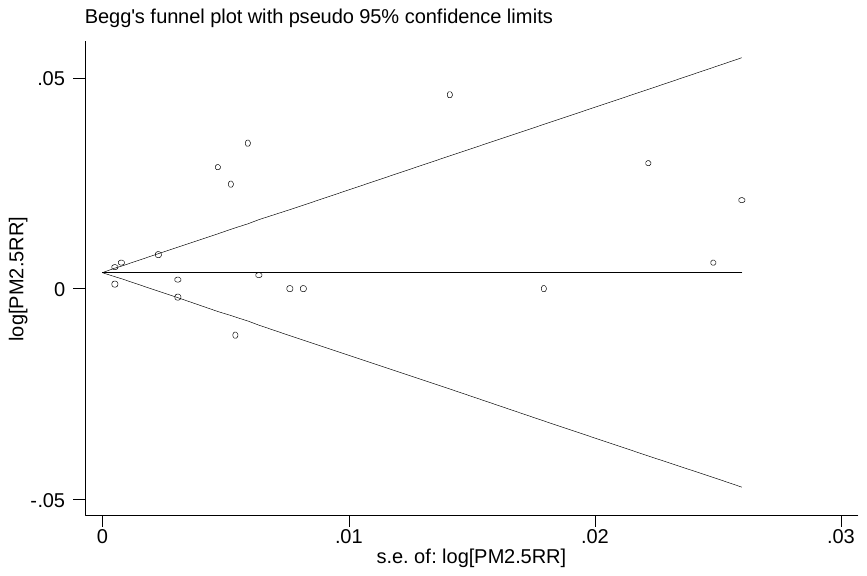

Figure S4-2 Begg’s funnel plot for relationship between PM_2.5_ and asthma exacerbations with lag0 exposure in quality sensitivity analyses (p=0.940)

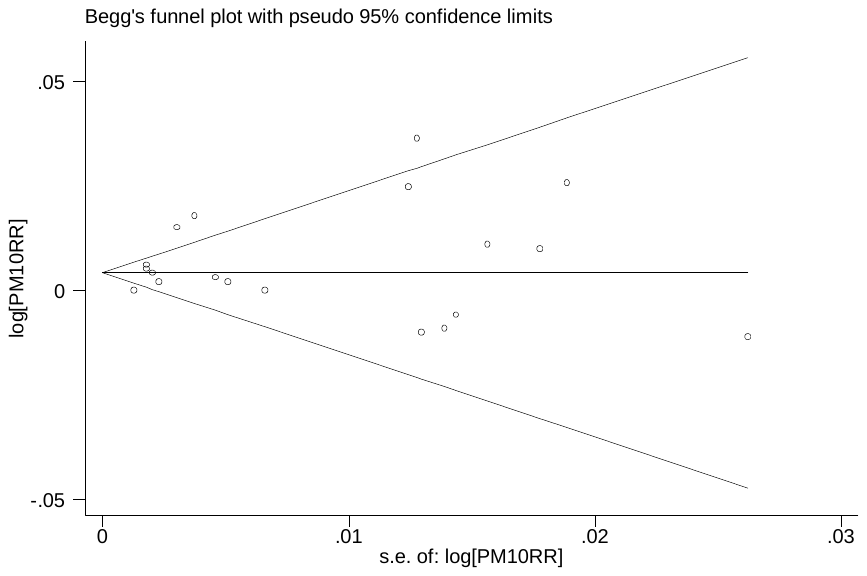

Figure S4-3 Begg’s funnel plot for relationship between PM_10_ and asthma exacerbations with lag0 exposure in quality sensitivity analyses (p=0.889)

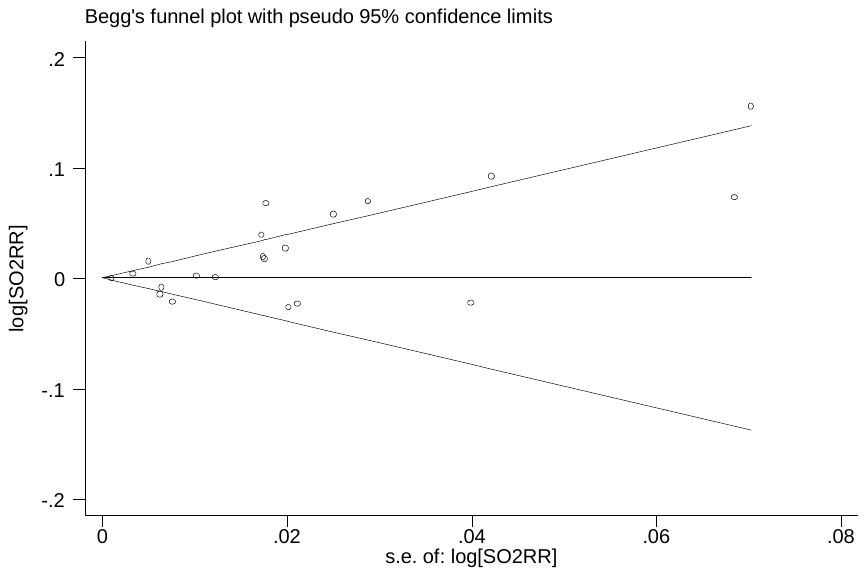

Figure S4-4 Begg’s funnel plot for relationship between SO_2_ and asthma exacerbations with lag0 exposure in quality sensitivity analyses (p=0.085)

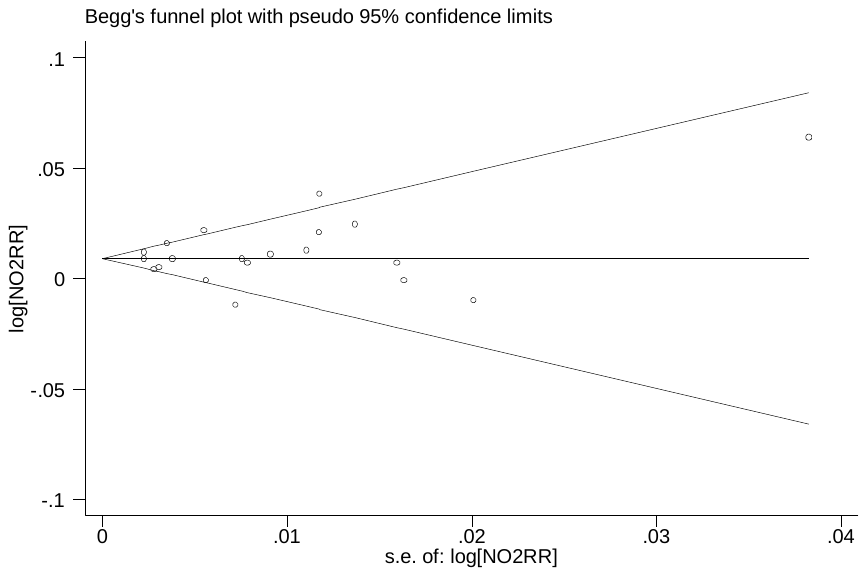

Figure 4-5 Begg’s funnel plot for relationship between NO_2_ and asthma exacerbations with lag0 exposure in quality sensitivity analyses (p=0.496)

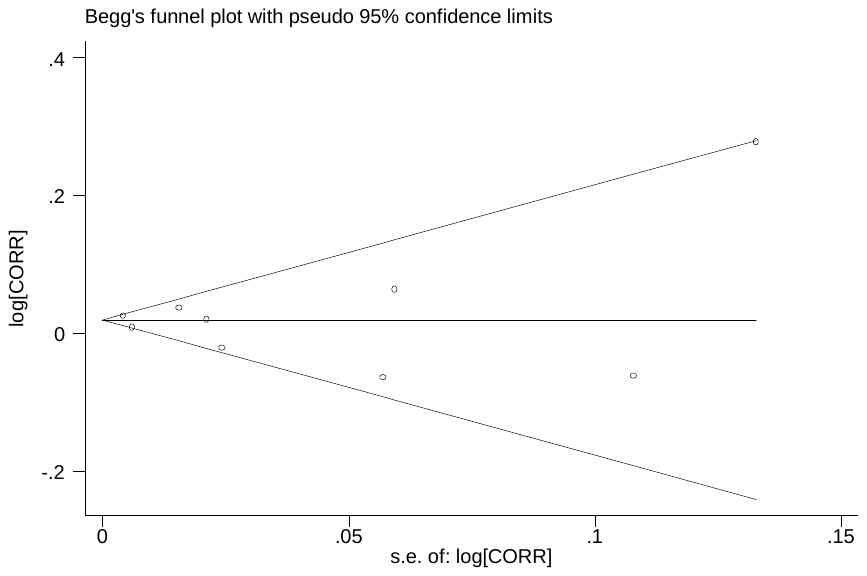

Figure 4-6 Begg’s funnel plot for relationship between CO and asthma exacerbations with lag0 exposure in quality sensitivity analyses (p=0.917)

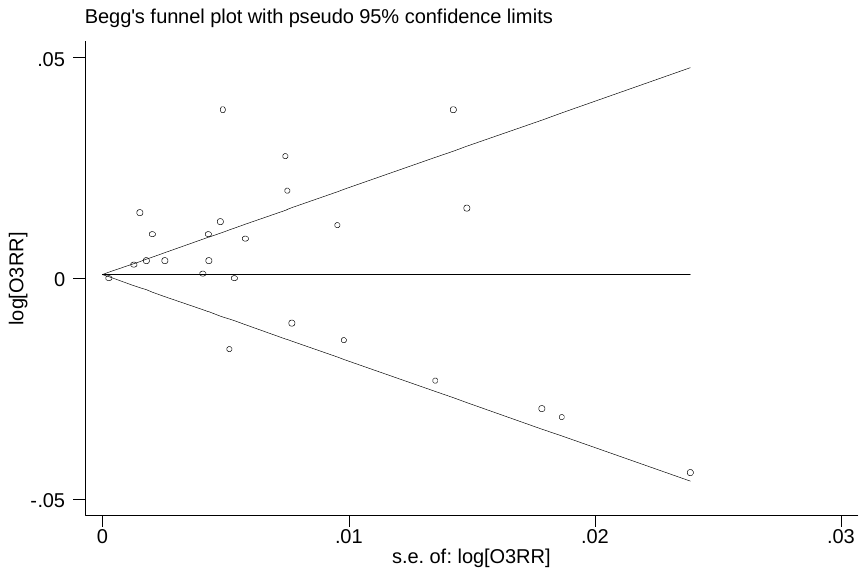

Figure 4-7 Begg’s funnel plot for relationship between O_3_ and asthma exacerbations with lag0 exposure in quality sensitivity analyses (p=0.047)

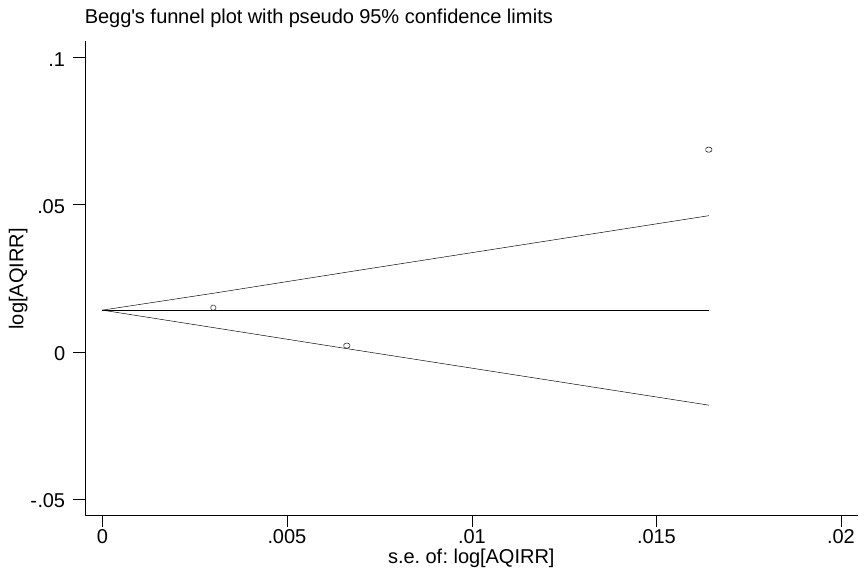

Figure 4-8 Begg’s funnel plot for relationship between AQI and asthma exacerbations with lag1 exposure in quality sensitivity analyses (p=1.000)

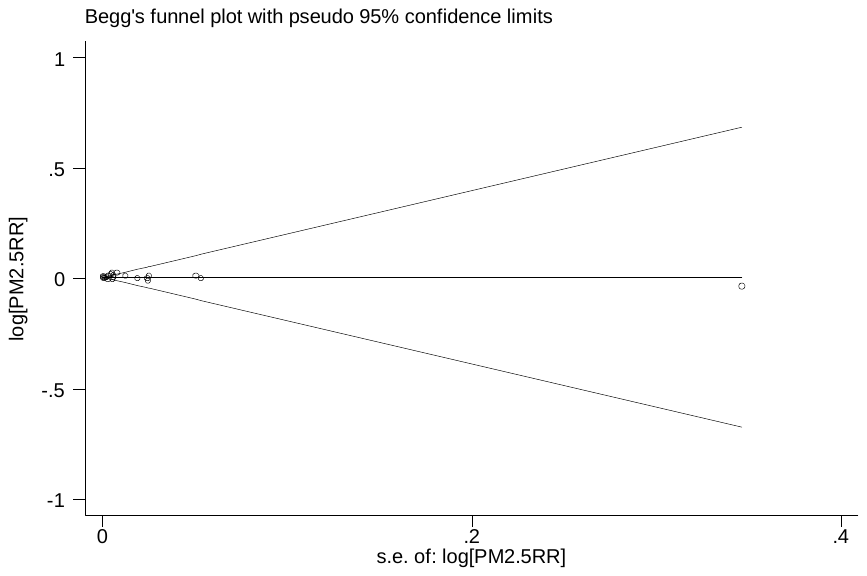

Figure 4-9 Begg’s funnel plot for relationship between PM_2.5_ and asthma exacerbations with lag1 exposure in quality sensitivity analyses (p=0.928)

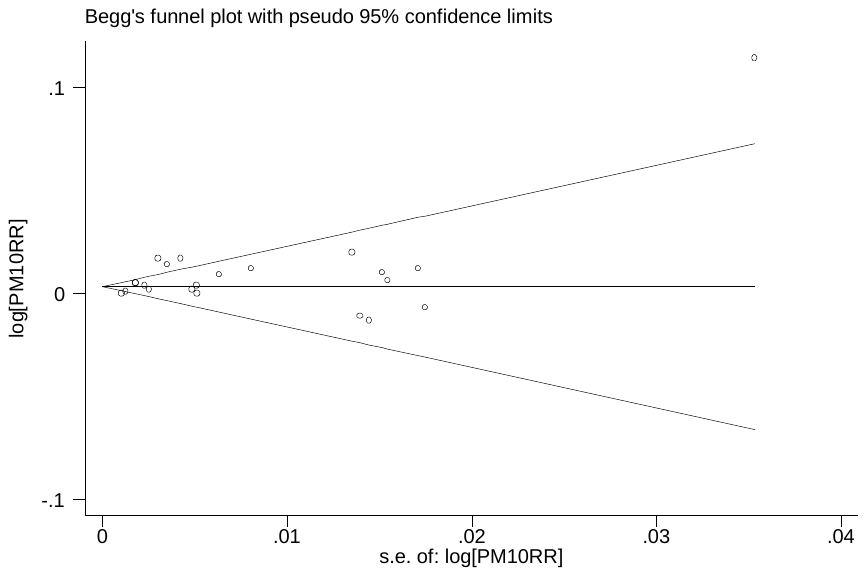

Figure 4-10 Begg’s funnel plot for relationship between PM_10_ and asthma exacerbations with lag1 exposure in quality sensitivity analyses (p=0.778)

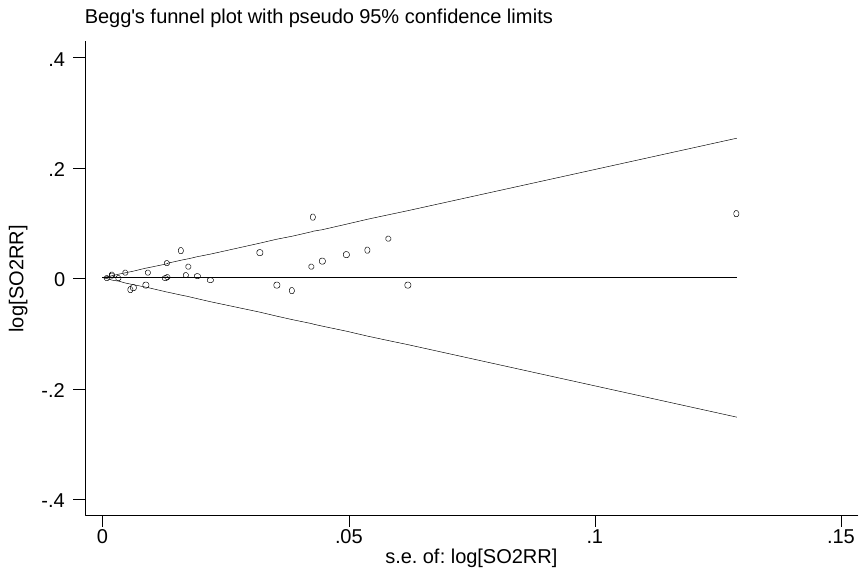

Figure 4-11 Begg’s funnel plot for relationship between SO_2_ and asthma exacerbations with lag1 exposure in quality sensitivity analyses (p=0.333)

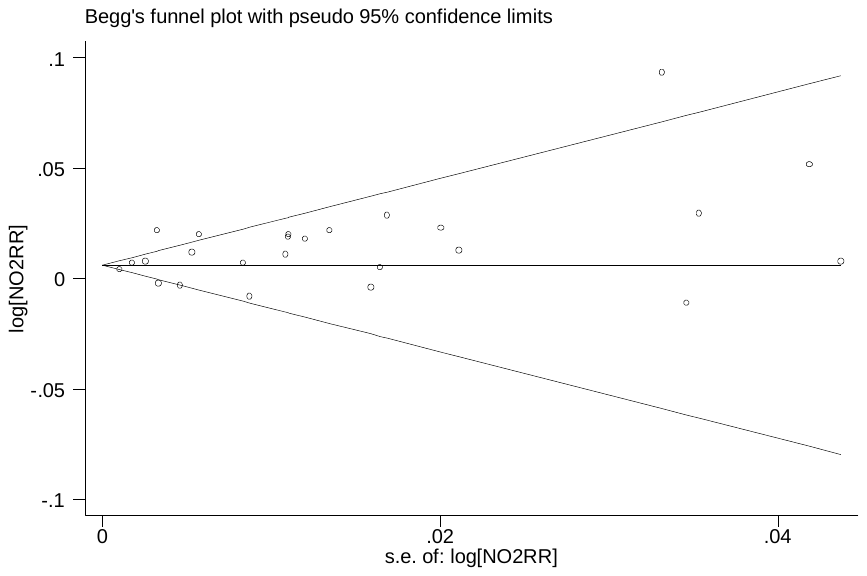

Figure 4-12 Begg’s funnel plot for relationship between NO_2_ and asthma exacerbations with lag1 exposure in quality sensitivity analyses (p=0.441)

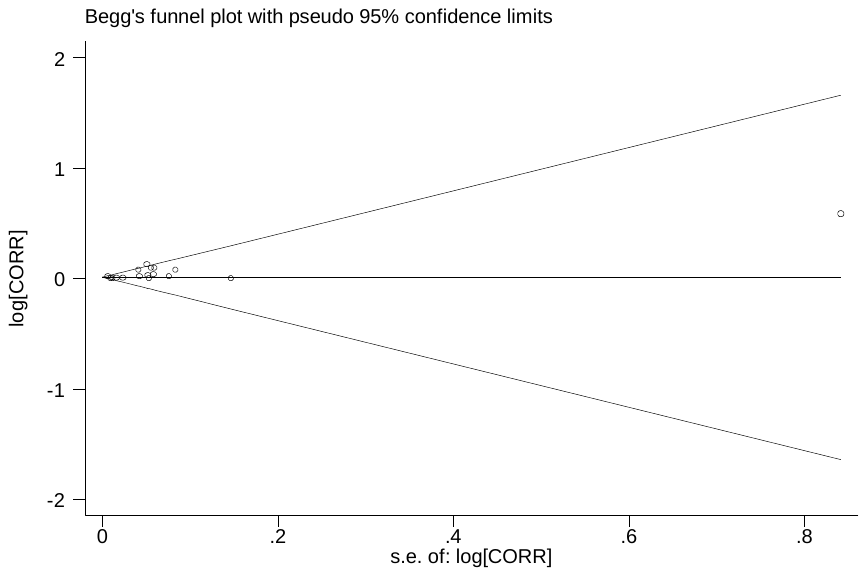

Figure 4-13 Begg’s funnel plot for relationship between CO and asthma exacerbations with lag1 exposure in quality sensitivity analyses (p=0.202)

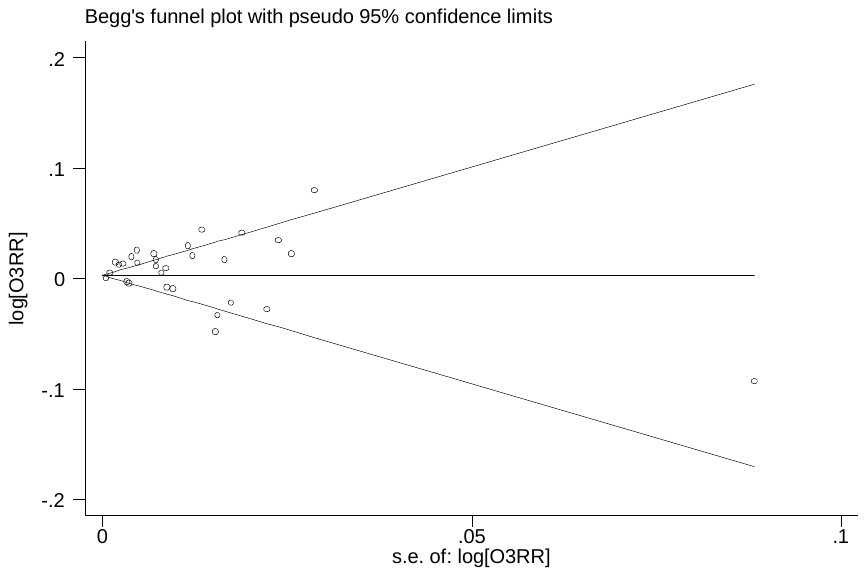

Figure 4-14 Begg’s funnel plot for relationship between O_3_ and asthma exacerbations with lag1 exposure in quality sensitivity analyses (p=0.116)
