## Supplementary material for "Outdoor air pollution and the risk of asthma exacerbations in single lag0 and lag1 exposure patterns: A systematic review and meta-analysis": Table S1. Search strategies

Database(s): Embase（693）、Web of Science (WOS)（1029）、Cochrane（64）、Pubmed（387）、ClinicalTrials（1）、China National Knowledge Internet (CNKI)（29）、Wanfang databases（61）、Chinese BioMedical (CBM)（10）

| **Embase** | **Search strategies** | **Results** |
| --- | --- | --- |
| 1 | 'asthma'/exp |  |
| 2 | 'bronchial asthma':ab,ti |  |
| 3 | 'asthma, bronchial':ab,ti |  |
| 4 | 'stridor*':ti,ab |  |
| 5 | 'asthma*':ab,ti |  |
| 6 | 'asthmatic':ab,ti |  |
| 7 | 'wheezing*':ab,ti |  |
| 8 | 'respiratory tract allergy':ab,ti |  |
| 9 | 'respiratory allergy':ab,ti |  |
| 10 | 'bronchospasm':ab,ti |  |
| 11 | 'bronchoconstriction':ab,ti |  |
| 12 | 'bronchial hyperreactivity':ab,ti |  |
| 13 | 'reactive airway disease':ab,ti |  |
| 14 | #1 OR #2 OR #3 OR #4 OR #5 OR #6 OR #7 OR #8 OR #9 OR #10 OR #11 OR #12 OR #13 |  |
| 15 | 'air pollution*':ab,ti |  |
| 16 | 'pollution, air':ab,ti |  |
| 17 | 'air quality':ab,ti |  |
| 18 | 'atmosphere pollution':ab,ti |  |
| 19 | 'atmospheric pollution':ab,ti |  |
| 20 | #15 OR #16 OR #17 OR #18 OR #19 |  |
| 21 | 'flareup*' |  |
| 22 | 'flareup*' |  |
| 23 | 'flare-up' |  |
| 24 | 'flaring up' |  |
| 25 | 'magnification*' |  |
| 26 | 'worsening' |  |
| 27 | 'acute symptom flare*' |  |
| 28 | 'acute exacerbation' |  |
| 29 | 'exacerbation*' |  |
| 30 | 'symptom increase' |  |
| 31 | 'increase,symptom' |  |
| 32 | 'symptom exaggeration*' |  |
| 33 | 'exaggeration,symptom' |  |
| 34 | #21 OR #22 OR #23 OR #24 OR #25 OR # 26 OR #27 OR #28 OR #29 OR #30 OR #31 OR #32 OR #33 |  |
| 35 | #14 AND #20 AND #34 |  |
| 36 | #14 AND #20 AND #34 AND [humans]/lim | 693 |

| **WOS** | **Search strategies** | **Results** |
| --- | --- | --- |
| 1 | TS=(asthma* OR Stridor* OR asthmatic OR wheezing* OR "respiratory tract allergy" OR "respiratory allergy" OR bronchospasm OR bronchoconstriction OR "bronchial hyperreactivity" OR "reactive airway disease" OR "Bronchial Asthma" OR "Asthma, Bronchial") |  |
| 2 | TS= ("Air Pollution*" OR "Pollution, Air" OR"Air Quality"OR "atmosphere pollution"OR"atmospheric pollution") |  |
| 3 | TS=("acute exacerbation" OR "Flare Up*" OR"Flareup*"OR "Flare-up"OR"Flaring Up" OR "Magnification*" OR"Worsening"OR "Acute Symptom Flare*"OR"Exacerbation*"OR "Symptom Increase" OR"Increase, Symptom" OR "Symptom Exaggeration*" OR "Exaggeration, Symptom") |  |
| 9 | #3 AND #2 AND #1 | 1029 |

| **Cochrane** | **Search strategies** | **Results** |
| --- | --- | --- |
| 1 | MeSH descriptor: [Asthma] explode all trees |  |
| 2 | (Stridor*): ti, ab, kw OR (asthmatic): ti, ab kw OR (wheezing*): ti, ab, kw OR (respiratory tract allergy): ti, ab, kw OR (respiratory allergy): ti, ab, kw (Word variations have been searched) |  |
| 3 | (bronchospasm): ti, ab, kw OR (bronchoconstriction): ti, ab kw OR (bronchial hyperreactivity): ti, ab, kw OR (reactive airway disease): ti, ab, kw OR (Bronchial Asthma): ti, ab, kw (Word variations have been searched) |  |
| 4 | (Asthma, Bronchial): ti, ab, kw (Word variations have been searched) |  |
| 5 | #1 or #2 or #3 or #4 |  |
| 6 | (Air Pollution*): ti, ab, kw OR (Pollution, Air): ti, ab kw OR (Air Quality): ti, ab, kw OR (atmosphere pollution): ti, ab, kw OR (atmospheric pollution): ti, ab, kw (Word variations have been searched) |  |
| 7 | (acute exacerbation) OR (Flare Up*) OR (Flareup*) OR (Flare-up) OR (Flaring Up) (Word variations have been searched) |  |
| 8 | (Magnification*) OR (Worsening) OR (Acute Symptom Flare*) OR (Exacerbation*) OR (Symptom Increase) (Word variations have been searched) |  |
| 9 | (Increase, Symptom) OR (Symptom Exaggeration*) OR (Exaggeration, Symptom) (Word variations have been searched) |  |
| 10 | #7 or #8 or #9 |  |
| 11 | #5 and #6 and #10 in Trials | 64 |

| **Pubmed** | **Search strategies** | **Results** |
| --- | --- | --- |
| 1 | Search AQI |  |
| 2 | Search "Asthma"[Mesh] |  |
| 3 | Search (((((((((((asthma*[Title/Abstract]) OR Stridor*[Title/Abstract]) OR asthmatic[Title/Abstract]) OR wheezing*[Title/Abstract]) OR respiratory tract allergy[Title/Abstract]) OR respiratory allergy[Title/Abstract]) OR bronchospasm[Title/Abstract]) OR bronchoconstriction[Title/Abstract]) OR bronchial hyperreactivity[Title/Abstract]) OR reactive airway disease[Title/Abstract]) OR Bronchial Asthma[Title/Abstract]) OR Asthma, Bronchial[Title/Abstract] |  |
| 4 | Search ("Asthma"[Mesh]) OR ((((((((((((asthma*[Title/Abstract]) OR Stridor*[Title/Abstract]) OR asthmatic[Title/Abstract]) OR wheezing*[Title/Abstract]) OR respiratory tract allergy[Title/Abstract]) OR respiratory allergy[Title/Abstract]) OR bronchospasm[Title/Abstract]) OR bronchoconstriction[Title/Abstract]) OR bronchial hyperreactivity[Title/Abstract]) OR reactive airway disease[Title/Abstract]) OR Bronchial Asthma[Title/Abstract]) OR Asthma, Bronchial[Title/Abstract]) |  |
| 5 | Search ((((Air Pollution*[Title/Abstract]) OR Pollution, Air[Title/Abstract]) OR Air Quality[Title/Abstract]) OR atmosphere pollution[Title/Abstract]) OR atmospheric pollution[Title/Abstract] |  |
| 6 | Search ((((((((((((acute exacerbation) OR Flare Up*) OR Flareup*) OR Flare-up) OR Flaring Up) OR Magnification*) OR Worsening) OR Acute Symptom Flare*) OR Exacerbation*) OR Symptom Increase) OR Increase, Symptom) OR Symptom Exaggeration*) OR Exaggeration, Symptom |  |
| 7 | Search (((("Asthma"[Mesh]) OR ((((((((((((asthma*[Title/Abstract]) OR Stridor*[Title/Abstract]) OR asthmatic[Title/Abstract]) OR wheezing*[Title/Abstract]) OR respiratory tract allergy[Title/Abstract]) OR respiratory allergy[Title/Abstract]) OR bronchospasm[Title/Abstract]) OR bronchoconstriction[Title/Abstract]) OR bronchial hyperreactivity[Title/Abstract]) OR reactive airway disease[Title/Abstract]) OR Bronchial Asthma[Title/Abstract]) OR Asthma, Bronchial[Title/Abstract]))) AND (((((Air Pollution*[Title/Abstract]) OR Pollution, Air[Title/Abstract]) OR Air Quality[Title/Abstract]) OR atmosphere pollution[Title/Abstract]) OR atmospheric pollution[Title/Abstract])) AND (((((((((((((acute exacerbation) OR Flare Up*) OR Flareup*) OR Flare-up) OR Flaring Up) OR Magnification*) OR Worsening) OR Acute Symptom Flare*) OR Exacerbation*) OR Symptom Increase) OR Increase, Symptom) OR Symptom Exaggeration*) OR Exaggeration, Symptom) |  |
| 8 | Search AQI Filters: Humans |  |
| 9 | Search (((("Asthma"[Mesh]) OR ((((((((((((asthma*[Title/Abstract]) OR Stridor*[Title/Abstract]) OR asthmatic[Title/Abstract]) OR wheezing*[Title/Abstract]) OR respiratory tract allergy[Title/Abstract]) OR respiratory allergy[Title/Abstract]) OR bronchospasm[Title/Abstract]) OR bronchoconstriction[Title/Abstract]) OR bronchial hyperreactivity[Title/Abstract]) OR reactive airway disease[Title/Abstract]) OR Bronchial Asthma[Title/Abstract]) OR Asthma, Bronchial[Title/Abstract]))) AND (((((Air Pollution*[Title/Abstract]) OR Pollution, Air[Title/Abstract]) OR Air Quality[Title/Abstract]) OR atmosphere pollution[Title/Abstract]) OR atmospheric pollution[Title/Abstract])) AND (((((((((((((acute exacerbation) OR Flare Up*) OR Flareup*) OR Flare-up) OR Flaring Up) OR Magnification*) OR Worsening) OR Acute Symptom Flare*) OR Exacerbation*) OR Symptom Increase) OR Increase, Symptom) OR Symptom Exaggeration*) OR Exaggeration, Symptom) Filters: Humans | 387 |

| **ClinicalTrials** | **Search strategies** | **Results** |
| --- | --- | --- |
| 1 | ( ( (题名=哮喘 或者v_subject=中英文扩展(哮喘,中英文对照) 或者(题名=气喘或者v_subject=中英文扩展(气喘,中英文对照)) ) 并且( (题名=空气污染 或者v_subject=中英文扩展(空气污染,中英文对照)) 或者(题名=大气污染或者v_subject=中英文扩展(大气污染,中英文对照) ) 并且( (题名=急性加重) 或者(题名=急性发作期) ) (模糊匹配) | 1 |

| **CNKI** | **Search strategies** | **Results** |
| --- | --- | --- |
| 1 | ( ( (主题=哮喘或者题名=哮喘或者v_subject=中英文扩展(哮喘,中英文对照) 或者title=中英文扩展(哮喘,中英文对照)) 或者(主题=气喘或者题名=气喘或者v_subject=中英文扩展(气喘,中英文对照) 或者title=中英文扩展(气喘,中英文对照)) ) 或者(主题=哮病或者题名=哮病或者v_subject=中英文扩展(哮病,中英文对照) 或者title=中英文扩展(哮病,中英文对照)) ) 并且( (主题=空气污染或者题名=空气污染或者v_subject=中英文扩展(空气污染,中英文对照) 或者title=中英文扩展(空气污染,中英文对照)) 或者(主题=大气污染或者题名=大气污染或者v_subject=中英文扩展(大气污染,中英文对照) 或者title=中英文扩展(大气污染,中英文对照)) ) 并且( (全文=急性加重) 或者(全文=急性发作期) ) (模糊匹配) | 29 |

| **Wanfang** | **Search strategies** | **Results** |
| --- | --- | --- |
| 1 | (主题:(哮喘+哮病+气喘)*主题:(大气污染+空气污染)*全部:(急性发作期+急性加重)) | 61 |

| **CBM** | **Search strategies** | **Results** |
| --- | --- | --- |
| 1 | "空气污染"[不加权:扩展] |  |
| 2 | 大气污染 |  |
| 3 | 急性加重 |  |
| 4 | 急性发作期 |  |
| 5 | 哮病 |  |
| 6 | 气喘 |  |
| 7 | (气喘) OR (哮病) OR ("哮喘"[不加权:扩展]) |  |
| 8 | (大气污染) OR ("空气污染"[不加权:扩展]) |  |
| 9 | (急性发作期) OR (急性加重) |  |
| 10 | ((急性发作期) OR (急性加重)) AND ((大气污染) OR ("空气污染"[不加权:扩展])) AND ((气喘) OR (哮病) OR ("哮喘"[不加权:扩展])) | 10 |
