## Supplementary material for "Outdoor air pollution and the risk of asthma exacerbations in single lag0 and lag1 exposure patterns: A systematic review and meta-analysis": Table S3. Relationships between air pollutants and asthma exacerbations in various outcomes and age subgroup analyses

| Table S3. Relationships between air pollutants and asthma exacerbations in various outcomes and age subgroup analyses | | | | | | | |
| --- | --- | --- | --- | --- | --- | --- | --- |
| Subgroup  (No.^a^, RR(95%CI) | **Air pollutants (incremental unit)** | | | | | | |
|  | **AQI (100units)** | **PM_2.5_ (10μg/m^3^)** | **PM_10_ (10μg/m^3^)** | **SO_2_ (10μg/m^3^)** | **NO_2_ (10μg/m^3^)** | **CO (1mg/m^3^)** | **O_3_ (10μg/m^3^)** |
| Age with lag0 exposure^b^ |  |  |  |  |  |  |  |
| Children | 2, 1.014(1.008, 1.020) | 6, 1.032(1.025, 1.039) | 7, 1.027(1.002, 1.052) | 9, 1.015(1.003, 1.027) | 7, 1.008(0.998, 1.018) | 6, 1.022(1.002, 1.041) | 11, 1.004(0.997, 1.011) |
| Adults | - | 4, 1.005(1.002, 1.008) | 4, 1.003(1.001, 1.004) | 7, 0.999(0.984, 1.013) | 6, 1.010(1.003, 1.018) | 3, 1.111(0.937, 1.286) | 7, 1.006(0.996, 1.017) |
| Age with lag1 exposure^c^ |  |  |  |  |  |  |  |
| Children | 2, 1.015(1.009, 1.021) | 7, 1.014(1.007, 1.021) | 7, 1.014(1.005, 1.023) | 11, 1.015(1.005, 1.025) | 7, 1.009(1.004, 1.014) | 8, 1.067(1.024, 1.110) | 12, 1.006(0.999, 1.012) |
| Adults | - | 2, 1.005(1.001, 1.009) | 2, 0.998(0.978, 1.019) | 3, 0.993(0.978, 1.009) | 3, 0.993(0.978, 1.009) | 1, 0.998(0.979, 1.017) | 6, 1.009(1.000, 1.019) |
| Outcomes with lag0 exposure^b^ |  |  |  |  |  |  |  |
| Outpatient visits | 2, 1.026(0.999, 1.054) | 4, 1.004(1.000, 1.007) | 2, 1.003(1.000, 1.006) | 1, 1.001(0.998, 1.005) | 3, 1.008(1.001, 1.015) | - | 2, 0.997(0.969, 1.024) |
| ER visits | - | 16, 1.010(1.003, 1.016) | 9, 1.017(0.998, 1.036) | 16, 1.004(1.000, 1.008) | 13, 1.012(1.002, 1.023) | 7, 1.028(0.992, 1.064) | 20, 1.009(1.004, 1.013) |
| Hospitalizations | 1, 0.998(0.976, 1.021) | 6, 1.008(1.002, 1.014) | 11, 1.004(1.002, 1.007) | 9, 1.005(0.992, 1.019) | 9, 1.009(1.005, 1.014) | 4, 1.021(0.997, 1.046) | 9, 1.004(1.001, 1.008) |
| ER or Hospitalizations | - | - | 1, 1.000(0.987, 1.013) | - | - | - | - |
| Deaths | - | 1, 1.003(0.990, 1.015) | 2, 1.003(0.996, 1.009) | 2, 0.992(0.967, 1.017) | 2, 1.017(0.996, 1.038) | 1, 1.066(0.941, 1.190) | 1, 0.986(0.967, 1.005) |
| Other events | 1, 1.201(1.155, 1.247) | 2, 1.047(1.024, 1.069) | 3, 1.119(1.018, 1.220) | 3, 1.029(0.982, 1.076) | 1, 1.033(1.000, 1.066) | 2, 1.124(1.051, 1.197) | 1, 0.913(0.877, 0.949) |
| Outcomes with lag1 exposure^c^ |  |  |  |  |  |  |  |
| Outpatient visits | 2, 1.040(0.986, 1.095) | 3, 1.007(0.999, 1.015) | 2, 1.003(0.991, 1.015) | 2, 1.012(0.993, 1.031) | 3, 1.006(0.998, 1.015) | - | 2, 1.006(0.983, 1.029) |
| ER visits | - | 14, 1.005(1.000, 1.011) | 7, 1.012(1.002, 1.022) | 11, 1.002(0.994, 1.009) | 12, 1.004(0.998, 1.009) | 9, 1.013(0.994, 1.032) | 17, 1.015(1.011, 1.020) |
| Hospitalizations | 1, 1.002(0.989, 1.015) | 7, 1.004(1.000, 1.008) | 12, 1.005(1.002, 1.008) | 13, 1.001(0.995, 1.007) | 9, 1.012(1.004, 1.020) | 5, 1.017(0.995, 1.039) | 13, 1.006(1.000, 1.012) |
| ER or Hospitalizations | - | - | 1, 1.009(0.996, 1.021) | - | - | - | - |
| Deaths | - | 1, 1.005(0.993, 1.017) | 2, 1.001(0.994, 1.008) | 2, 1.002(0.981, 1.024) | 2, 1.015(0.994, 1.036) | 1, 1.035(0.917, 1.153) | 1, 0.992(0.975, 1.009) |
| Other events | 1, 1.204(1.158, 1.251) | 1, 1.080(1.010, 1.150) | 3, 1.122(1.015, 1.230) | 3, 1.034(0.968, 1.099) | 1, 1.046(1.013, 1.079) | 2, 1.155(1.037, 1.274) | 2, 0.985(0.938, 1.032) |
| Abbreviations: PM_2.5_, particulate matter diameter ≤ 2.5μ m; PM_10_, particulate matter diameter ≤ 2.5μ m; RR, relative risk.  ^a^ No., No. of the studies.  ^b^ lag0 exposure, single lag0 exposure.  ^c^ lag1 exposure, single lag1 exposure. | | | | | | | |
